# Meditation Practice and Long-Term Mortality in US Adults: A Doubly Robust Analysis of Pooled National Health Interview Survey Cohorts with a Trial-Informed Bayesian Subgroup Analysis

**DOI:** 10.64898/2026.09.17.26363306

**Authors:** Lu Shi, Yian Gu, Hafsa Imtiaz, Donglan S. Zhang, Anthony D. Mancini

## Abstract

**Introduction:** Trial evidence in older adults with hypertension suggests that meditation may lower mortality, but this association has not been tested in a nationally representative study with linked-mortality data. This study estimated meditation–mortality associations among US adults, examined variation by family income, and synthesized trial and survey evidence for the population matched to the trial evidence base.

**Methods:** We pooled four National Health Interview Survey cohorts (2002–2017) with mortality follow-up through 2019 (N=90,041; 8,402 [9.3%] reported meditation practice in the past 12 months). Doubly robust survey-weighted Cox models combined stabilized inverse-probability weights with covariate adjustment, overall and by income stratum; 15-year risks were standardized using g-computation, and probabilistic bias analysis addressed exposure misclassification, including a differential-by-income scenario. For non-Hispanic Black adults ≥55 years with hypertension (n=2,786), Bayesian synthesis combined subgroup estimates with a pooled randomized-trial prior.

**Results:** Over 818,248 person-years of follow-up, 10,027 all-cause and 3,040 cardiovascular deaths occurred. Doubly robust hazard ratios (HR) were 1.02 (95% CI 0.90–1.16) for all-cause and 0.96 (95% CI 0.79–1.16) for cardiovascular mortality. Among individuals below 200% of the federal poverty level, HRs were 0.85 (All-cause: 95% CI 0.71–1.03) and 0.80 (Cardiovascular mortality: 95% CI 0.57–1.10), compared with 1.10 (95% CI 0.93–1.30) and 1.02 (95% CI 0.80–1.30) among those at/above that threshold (interaction p=0.19 and p=0.62). In the subgroup analysis, posteriors under a 50%-discounted trial prior were 0.84 (All-cause mortality: Pr[HR<1]=0.89) and 0.70 (Cardiovascular mortality: Pr[HR<1]=0.96). **Conclusions:** Meditation practice showed no overall association with mortality after doubly robust adjustment. However, estimates suggesting a protective effect were concentrated among adults below 200% of the federal poverty level, and trial-informed synthesis indicated probable benefit in cardiovascular mortality in the high-risk subgroup most comparable to trial participants. These hypothesis-generating findings motivate further study of effect modification using more detailed exposure measurement.

## INTRODUCTION

Meditation practice has grown substantially among US adults, with prevalence rising from 7.5% in 2002 to 17.3% in 2022.^1^ Randomized trials indicate that meditation can modestly lower blood pressure,^2^ the American Heart Association concluded that meditation may be considered as an adjunct to guideline-directed cardiovascular risk reduction while emphasizing the limited quality and quantity of the underlying evidence,^3^ and the Mindfulness-Based Blood Pressure Reduction trial has since demonstrated clinically meaningful systolic reductions in adults with elevated office blood pressure.^4^ For mortality, the most direct evidence is a pooled analysis of two randomized trials of Transcendental Meditation in persons aged ≥55 years with systemic hypertension, which reported a relative risk of 0.77 (p=0.039) for all-cause and 0.70 (p=0.045) for cardiovascular mortality.^5^ To our knowledge, however, no nationally representative linked-mortality analysis has tested whether meditation practice is associated with subsequent mortality, or whether any such association differs by economic circumstances, given that economic adversity is associated with higher mortality risk and may shape the value of stress-reducing practices.

We used a two-tier design that assigns each analytic tool to the sample where it has power. In the full pooled National Health Interview Survey (NHIS) sample, doubly robust models estimated overall associations and income-stratified associations and their interaction. In the subgroup matched to the trial evidence base (non-Hispanic Black adults aged ≥55 years with hypertension), where sparse events preclude precise frequentist estimation, Bayesian synthesis combined the survey estimate with the trial prior. Because the exposure is self-reported, we quantified the potential impact of misclassification throughout.

## METHODS

### Study population

The NHIS is an annual household survey of the civilian, noninstitutionalized US population. The 2002, 2007, 2012, and 2017 adult samples included meditation items and are linked to National Death Index records through December 31, 2019, in the public-use Linked Mortality Files.^6,7^ We pooled linkage-eligible adults with complete data on the exposure and covariates (N=90,041). The pre-specified subgroup comprised non-Hispanic Black respondents aged ≥55 years reporting a hypertension history (n=2,786; 198 practiced meditation).

### Exposure

Meditation items were drawn from the NHIS complementary health questionnaires, whose cognitive testing documented heterogeneity in the practices respondents reported as meditation.^8^ In 2002, 2007, and 2012, meditation was ascertained within relaxation-technique items (ever use; past-12-month use); in 2017, respondents were asked about past-12-month practice of spiritual, mindfulness, or mantra meditation. The primary exposure was any past-12-month practice (n=8,402, 9.3%). A three-level recency proxy (never / past use only / past-12-month use), derived from survey items available in 2002–2012 (n=67,409), served as a secondary exposure.

### Outcomes

Outcomes were all-cause mortality and cardiovascular mortality (underlying cause in the public-use leading-cause groups “diseases of heart” or “cerebrovascular diseases” — the standard public-use cardiovascular definition in pooled NHIS-LMF cohort analyses^9^), from interview to death or December 31, 2019.

### Full-sample analysis

We plotted crude Kaplan–Meier survival by practice status within income strata (below vs at/above 200% of the federal poverty level [FPL]). Propensity scores from logistic regression (age and its square, sex, race/ethnicity, marital status, smoking status, hypertension, body mass index, region, income stratum, diabetes, an indicator of activity limitation attributed to depression or anxiety, and survey wave) yielded stabilized inverse-probability weights, truncated at the 1st and 99th percentiles^10^ and combined with sampling weights,^11^ in design-based Cox models^12^ (pooled strata and primary sampling units, wave-stratified baseline hazards) that additionally adjusted for the same covariates — a doubly robust specification.^13^ Because hypertension, diabetes, body mass index, smoking, and the depression indicator may lie partly downstream of sustained practice, a companion minimal-adjustment model (age, sex, race/ethnicity, marital status, region, income, wave) estimated the total association without conditioning on potential mediators.^14^ Income-stratified models re-estimated propensity scores within stratum, and a practice-by-income product term tested multiplicative effect-measure modification, with stratum-specific estimates reported alongside the interaction term.^15^

Because hazard ratios are not stand-alone causal effect measures,^16^ we standardized 15-year risks by g-computation^17^ from a weighted pooled logistic model on annual person-period data (natural-spline time-by-exposure interaction plus covariates), with counterfactual survival computed over a full 15-year grid, averaged over the 2002 and 2007 cohorts (whose follow-up spans the window; n=39,632), and bootstrapped by assigning exponential multiplier weights to primary sampling units (300 replicates). Probabilistic bias analysis addressed exposure misclassification.^18,19^ For nondifferential misclassification, sensitivity and specificity were sampled from U(0.70, 0.95) and U(0.85, 0.99) over 300 iterations; draws yielding inadmissible corrected prevalences were discarded. In each iteration, exposure status was reassigned using stratum-specific predictive values, weights were re-estimated, and the doubly robust model was refit. A differential scenario allowed greater over-reporting at higher incomes (specificity U[0.85, 0.97] at/above vs U[0.90, 0.99] below 200% FPL) and targeted the interaction estimate. E-values quantified sensitivity to unmeasured confounding.^20^

### Trial-informed Bayesian subgroup analysis

For the subgroup analysis, the doubly robust log hazard ratio and its variance provided the likelihood in a conjugate normal–normal update. The primary prior was centered on the pooled trial estimates (relative risks 0.77 and 0.70),^5^ with variance derived from the reported p-values. Prior precision was discounted by 50% because NHIS measures heterogeneous self-reported meditation practice rather than randomized Transcendental Meditation. Sensitivity analyses considered both the undiscounted trial prior and a skeptical prior (mean 0, SD 0.35 on the log scale). We report posterior hazard ratios, 95% credible intervals, and posterior probabilities of benefit. Analyses were conducted in R (survey and survival packages); all data were publicly available and de-identified.

## RESULTS

Of 90,041 adults, 8,402 (9.3%) reported meditation practice in the past 12 months; 31,749 (35.3%) had family income below 200% FPL (2,461 practitioners below and 5,941 at/above). Over 818,248 person-years (median follow-up 7.75 years), 10,027 all-cause and 3,040 cardiovascular deaths occurred (Table 1). Propensity-score weighting balanced all substantive covariates (standardized differences ≤0.07, from ≤0.43 unweighted); survey-wave indicators retained differences up to 0.16 and were additionally controlled by wave-stratified baseline hazards.

**Table 1.**
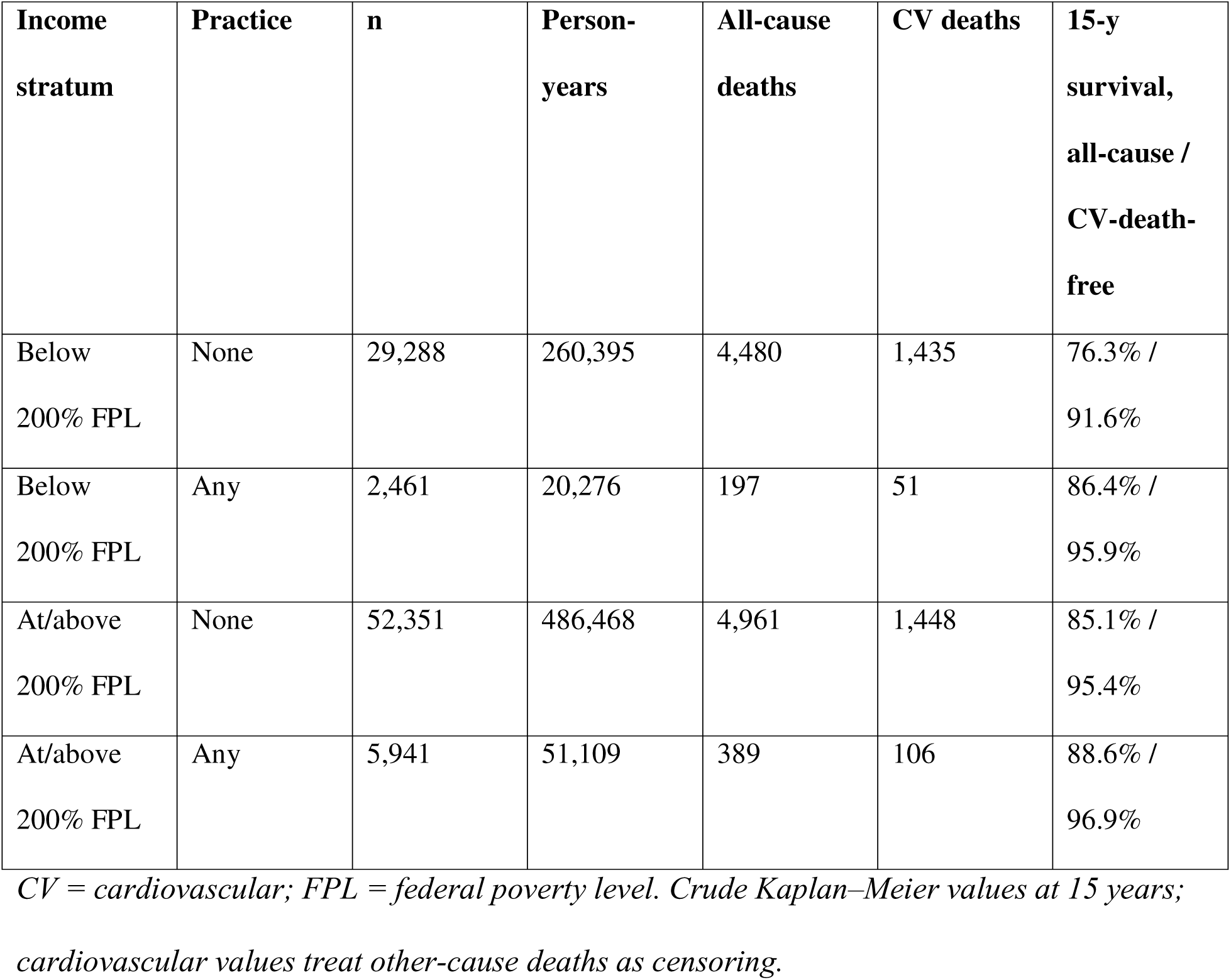
Sample, Person-Years, Deaths, and Crude 15-Year Survival by Meditation Practice and Income Stratum (N=90,041)

| **Income stratum** | **Practice** | **n** | **Person-years** | **All-cause deaths** | **CV deaths** | **15-y survival, all-cause / CV-death-free** |
| --- | --- | --- | --- | --- | --- | --- |
| Below 200% FPL | None | 29,288 | 260,395 | 4,480 | 1,435 | 76.3% / 91.6% |
| Below 200% FPL | Any | 2,461 | 20,276 | 197 | 51 | 86.4% / 95.9% |
| At/above 200% FPL | None | 52,351 | 486,468 | 4,961 | 1,448 | 85.1% / 95.4% |
| At/above 200% FPL | Any | 5,941 | 51,109 | 389 | 106 | 88.6% / 96.9% |

*CV = cardiovascular; FPL = federal poverty level. Crude Kaplan–Meier values at 15 years;* *cardiovascular values treat other-cause deaths as censoring.*

Crude survival analyses suggested better outcomes among meditation practitioners in both income strata, with larger differences among those below 200% FPL (Figure 1): 15-year all-cause survival was 10.1 percentage points higher among those below compared with 3.5 percentage points higher among those at or above 200% FPL. Corresponding differences for cardiovascular mortality were 4.2 and 1.5 percentage points.

**Figure 1.**
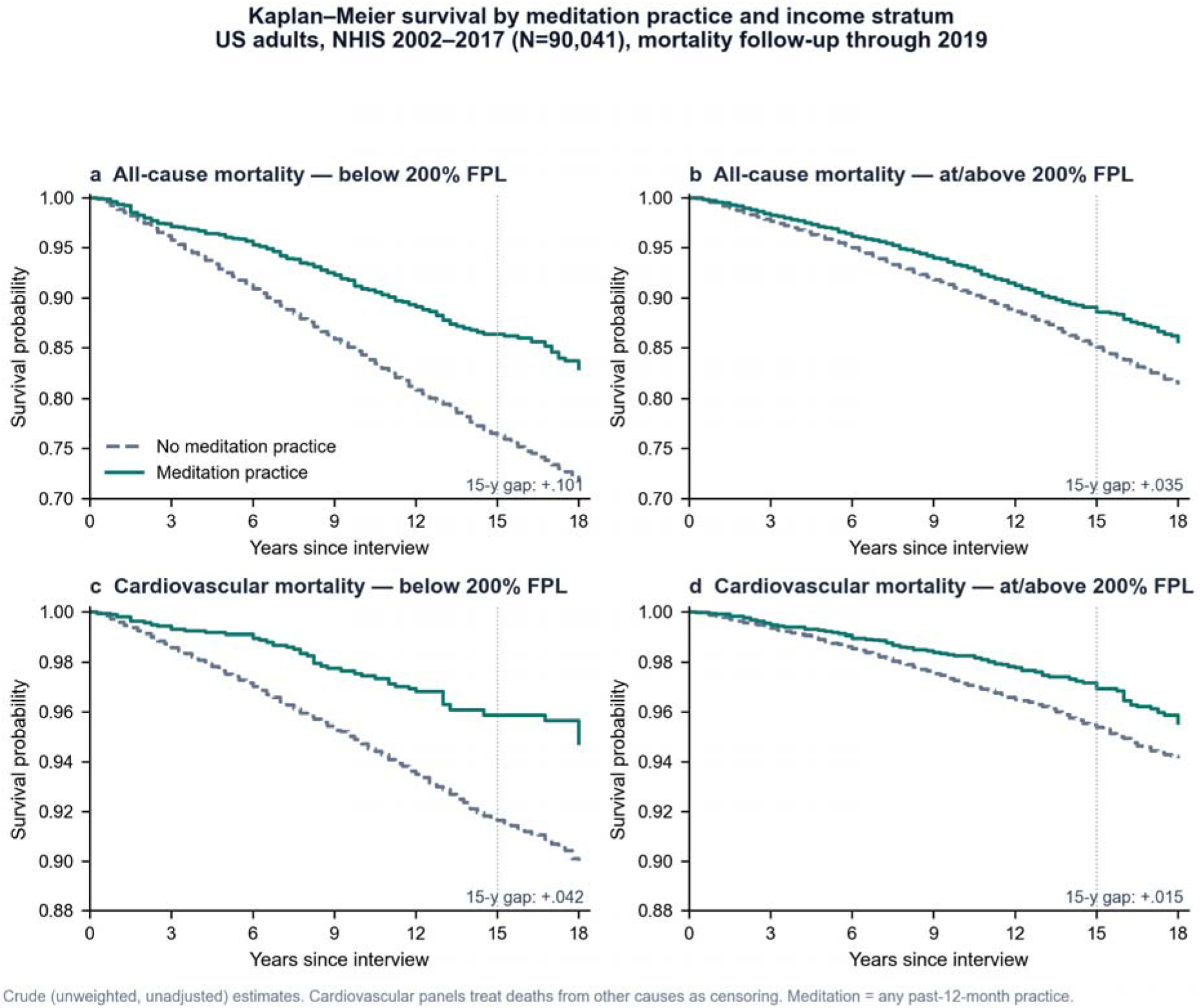
Kaplan–Meier Survival by Meditation Practice and Income Stratum. *Crude (unweighted, unadjusted) estimates, N=90,041. Meditation = any past-12-month practice. Cardiovascular panels treat deaths from other causes as censoring*.

These differences largely disappeared after adjustment (Table 2). Doubly robust hazard ratios were 1.02 (95% CI 0.90–1.16) for all-cause mortality and 0.96 (95% CI 0.79–1.16) for cardiovascular mortality, similar to covariate-adjusted estimates (0.98 [95% CI 0.86–1.12] and 0.94 [95% CI 0.78–1.14]) and minimally adjusted total-association models (1.03 [95% CI 0.91– 1.17] and 0.98 [95% CI 0.81–1.19]). This consistency suggests that the overall null findings were not driven by adjustment for potential mediators. In contrast, weighting-only models (0.82 [95% CI 0.72–0.93] and 0.72 [95% CI 0.59–0.88]) continued to suggest lower mortality, indicating residual confounding that was addressed by additional outcome-model adjustment. Standardized 15-year risks were 14.2% under meditation practice versus 14.7% under no practice for all-cause mortality (difference –0.4 percentage points, 95% CI –1.8 to +1.3) and 5.5% versus 5.8% for cardiovascular mortality (–0.2, –1.5 to +1.0).

**Table 2.** Hazard Ratios for Any Meditation Practice: Full-Sample Models and Trial-Informed Subgroup Synthesis.

| Model | All-cause mortality, HR (95% CI) | Cardiovascular mortality, HR (95% CI) |
| --- | --- | --- |
| **Full sample (N=90,041)** |  |  |
| Covariate-adjusted (survey Cox) | 0.98 (95% CI 0.86–1.12) | 0.94 (95% CI 0.78–1.14) |
| Inverse-probability-weighted only | 0.82 (95% CI 0.72–0.93) | 0.72 (95% CI 0.59–0.88) |
| Doubly robust (weights + adjustment) | 1.02 (95% CI 0.90–1.16) | 0.96 (95% CI 0.79–1.16) |
| Doubly robust, minimal adjustment (total association) | 1.03 (95% CI 0.91–1.17) | 0.98 (95% CI 0.81–1.19) |
| Doubly robust, below 200% FPL | 0.85 (95% CI 0.71–1.03) | 0.80 (95% CI 0.57–1.10) |
| Doubly robust, at/above 200% FPL | 1.10 (95% CI 0.93–1.30) | 1.02 (95% CI 0.80–1.30) |
| Ratio of HRs (below vs at/above) | 0.85 (95% CI 0.66–1.09);<br>p=0.19 | 0.90 (95% CI 0.59–1.37);<br>p=0.62 |
| QBA-corrected, median (95% simulation interval) | 1.02 (0.91–1.13) | 0.97 (0.81–1.16) |
| Interaction ratio under differential QBA scenario | 0.91 (0.58–1.56) | 0.95 (0.43–2.16) |
| **Subgroup: non-Hispanic Black, ≥55 y, hypertension** |  |  |
| **(n=2,786)** |  |  |
| Doubly robust (frequentist) | 0.97 (95% CI 0.63–1.49) | 0.70 (95% CI 0.36–1.38) |
| Posterior, trial prior | 0.81 (0.66–1.01); Pr=0.97 | 0.70 (0.51–0.96); Pr=0.99 |
| Posterior, 50%-discounted trial prior (primary) | 0.84 (0.64–1.11); Pr=0.89 | 0.70 (0.47–1.04); Pr=0.96 |
| Posterior, skeptical prior | 0.97 (0.68–1.41); Pr=0.55 | 0.84 (0.52–1.35); Pr=0.77 |

*HR = hazard ratio; CI = confidence interval (credible interval for posteriors); FPL = federal poverty level; QBA = probabilistic quantitative misclassification analysis (nondifferential: sensitivity 0.70–0.95, specificity 0.85–0.99; differential scenario: specificity 0.85–0.97 at/above vs 0.90–0.99 below 200% FPL); Pr = posterior probability HR<1. Minimal adjustment conditions only on age, sex, race/ethnicity, marital status, region, income, and wave. Trial prior from the pooled randomized trials of Transcendental Meditation in persons ≥55 years with systemic hypertension (reference 5); the primary prior discounts trial precision by 50% for exposure-mismatch transportability.*

Income-stratified doubly robust estimates differed in direction. Among adults below 200% FPL, hazard ratios were 0.85 (95% CI 0.71–1.03; p=0.09) for all-cause mortality and 0.80 (95% CI 0.57–1.10) for cardiovascular mortality, compared with 1.10 (95% CI 0.93–1.30) and 1.02 (95% CI 0.80–1.30), respectively, among those at or above 200% FPL. Corresponding ratios of hazard ratios were 0.85 (95% CI 0.66–1.09) (p=0.19) for all-cause mortality and 0.90 (95% CI 0.59– 1.37) (p=0.62) for cardiovascular mortality. In recency analyses, past meditation use was associated with lower all-cause mortality (HR 0.81, 95% CI 0.68–0.96), whereas current use was not (HR 0.97, 95% CI 0.84–1.12); corresponding cardiovascular mortality estimates were 0.86 (95% CI 0.64–1.15) and 0.92 (95% CI 0.75–1.12).

Results were largely unchanged after correcting for exposure misclassification. Median corrected doubly robust hazard ratios were 1.02 (95% simulation interval 0.91–1.13) for all-cause mortality and 0.97 (0.81–1.16) for cardiovascular mortality; incorporating random error widened these ranges to 0.84–1.26 and 0.67–1.30, respectively. Under the differential-by-income scenario, median corrected interaction ratios were 0.91 (0.58–1.56) for all-cause mortality and 0.95 (0.43–2.16) for cardiovascular mortality. E-values were 1.15 and 1.21 for the overall estimates, and 1.48 and 1.62 for the estimates among adults below 200%-FPL.

In the trial-matched subgroup, doubly robust hazard ratios were imprecise (0.97 [95% CI 0.63– 1.49] for all-cause mortality and 0.70 [95% CI 0.36–1.38] for cardiovascular mortality). Incorporating prior evidence from randomized trials shifted the posterior estimates toward benefit. Under the 50%-discounted trial prior, posteriors hazard ratios were 0.84 (95% credible interval 0.64–1.11; Pr[HR<1]=0.89) for all-cause mortality and 0.70 [95% CI 0.47–1.04; Pr[HR<1]=0.96] for cardiovascular mortality (Figure 2). The undiscounted trial prior yielded corresponding probabilities of benefit of 0.97 and 0.99, while the skeptical prior yielded probabilities of 0.55 and 0.77.

**Figure 2.**
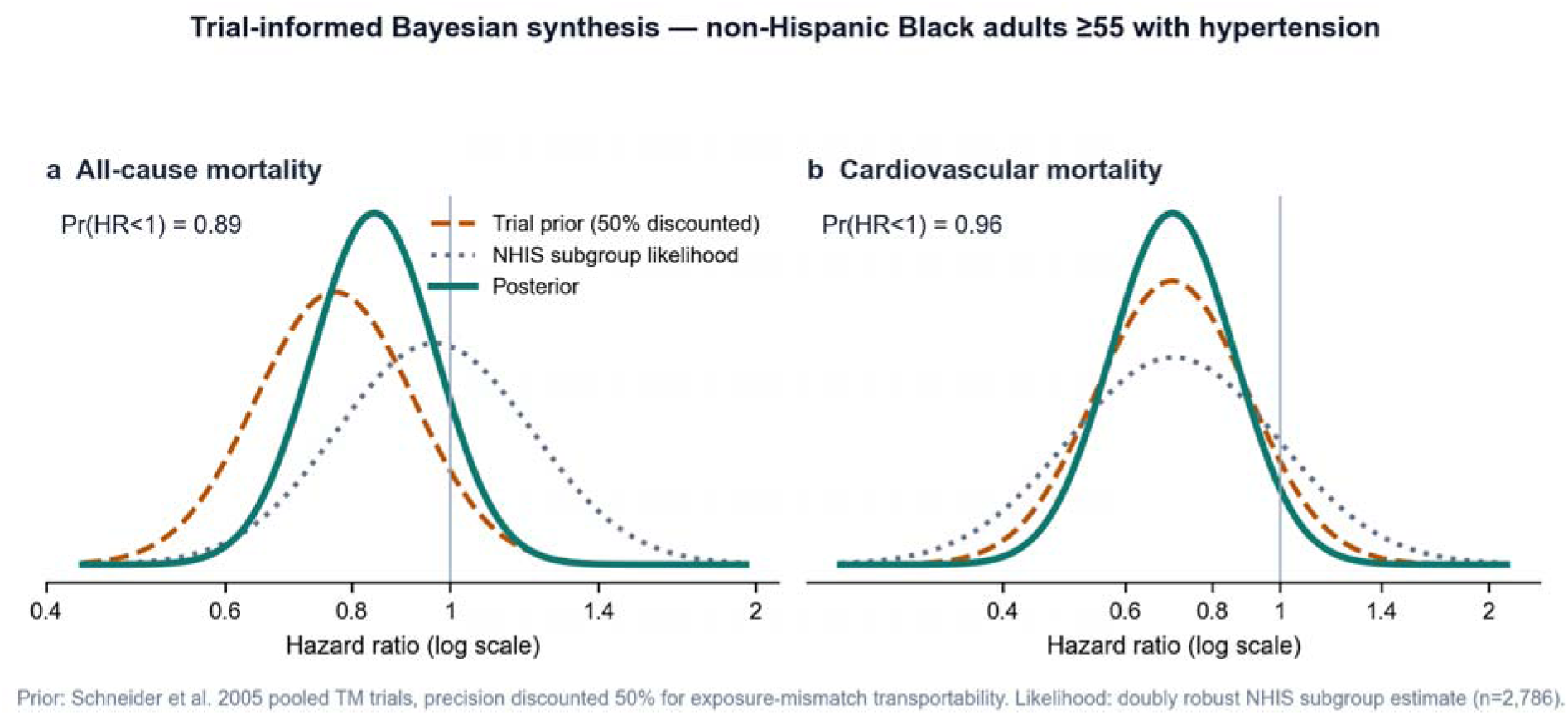
Trial-Informed Bayesian Synthesis in the Subgroup Matched to the Trial Evidence Base. Prior: pooled randomized trials of Transcendental Meditation (reference 5), precision discounted 50%. Likelihood: doubly robust NHIS estimate among non-Hispanic Black adults ≥55 years with hypertension (n=2,786). Posterior probabilities of benefit under undiscounted and skeptical priors appear in Table 2.

## DISCUSSION

In this national analysis powered by 90,041 adults and more than 10,000 deaths, meditation practice showed no overall association with long-term mortality after doubly robust adjustment. Similar findings from and minimal adjustment models suggest that this null association was not driven by adjustment for potential mediators. The large crude survival advantages reflected who takes up practice, not what practice does on average. Two patterns persisted after adjustment. First, estimates in the protective direction were concentrated among adults below 200% FPL for both outcomes, whereas hazard ratios were near or above one among those with higher incomes, although tests of interaction did not reach conventional level of statistical significance. Second, in the high-risk subgroup matched to the randomized trial population, Bayesian synthesis with trial-informed priors assigned a high posterior probability to cardiovascular benefit.

The two analytic tiers serve distinct but are complementary purposes. The full sample provides the precision needed to evaluate potential effect modification, with income-stratified confidence intervals that were roughly four times narrower than would have been possible in the subgroup alone. The subgroup analysis, in contrast, places the survey findings alongside the only randomized evidence on mortality,^5^ making transparent how conclusions change as different levels of confidence are assigned to that evidence through the prior. The posterior probabilities of cardiovascular benefit ranged from 0.99 under full confidence in the trial evidence to 0.77 under a skeptical prior, illustrating both the promise and the uncertainty of the combined evidence. The consistently favorable cardiovascular estimates are consistent with meta-analytic blood-pressure effects,^2^ and population-level modeling suggests that even modest reductions in cardiovascular risk, if causal, could translate into meaningful numbers of prevented cardiovascular events nationwide.^21^

The income-related pattern can be interpreted in at least two ways. One possibility is that higher baseline mortality at lower income levels yields larger absolute benefits even if relative effects are constant. Alternatively, stress-buffering mechanisms may confer greater relative benefits among adults facing greater cumulative physiological burden of adversity, consistent with the “weathering” hypothesis described in the allostatic-load literature.^22^ Stratified hazard ratios diverging in direction lean toward the latter explanation, and the differential-misclassification analysis indicates that income-related differences in over-reporting alone are unlikely to account for the observed pattern. However, interactions were assessed on the multiplicative scale only and were imprecise. Cohorts with repeated, richer measurement on meditation practices are needed to formally evaluate effect modification.

This analysis has several limitations. Meditation practice was self-reported at a single interview without information on duration or intensity, and survey wording differed in 2017. The three-level exposure measure captures recency rather than dose, and the modestly protective estimate for past-only use cautions against causal interpretations of any single comparison. Bias analysis addressed misclassification under stated parameter ranges but could not account for changes in practice over time. Educational attainment was unavailable in the analytic file, so socioeconomic adjustment relied on income, region, and marital status; the modest E-values suggest that relatively weak unmeasured confounding could explain the estimates observed below 200% FPL. Public-use mortality files perturb selected follow-up dates and causes of death, although National Center for Health Statistics evaluations indicate that resulting hazard ratios are generally similar, with only minor attenuation.^23^ Cardiovascular analyses treated deaths from other causes as censoring events. Income was measured only once, and Bayesian results necessarily depend on assumptions about the transportability of the trial evidence, as reflected in the prior-discounting strategy.

Within these limits, the study contributes design-consistent national estimates of meditation– mortality associations with explicit quantification of misclassification uncertainty, a powered test of income patterning, and a transparent synthesis with the randomized evidence. The results support further investigation into whether low-cost, self-administered contemplative practices may yield the largest benefits among populations facing the greatest economic adversity.

## Data Availability

All data used are available online at National Center for Health Statistics https://www.cdc.gov/nchs/linked-data/mortality-files/index.html

## ACKNOWLEDGMENTS

Author contributions: L.S. designed the study, conducted the statistical analyses, and drafted the manuscript; H.I. contributed to the statistical analyses and manuscript preparation; Y.G., D.S.Z. and A.D.M. contributed to interpretation and critical revision of the manuscript. No financial disclosures were reported by the authors of this paper. No specific funding supported this analysis. NHIS and public-use Linked Mortality Files are freely available from the National Center for Health Statistics; the complete replication R programs, which build the analytic files directly from the federal source files, appear in the Appendix.

## APPENDIX. Replication R Programs

This appendix contains the complete R programs used for the analyses in this article, in run order (A1–A7). Program A1 constructs the pooled analytic files directly and transparently from the federal source files: the NHIS 2002, 2007, and 2012 Sample Adult (SAMADULT), Sample Adult Alternative Medicine (ALTHEALT), and Family (FAMILYXX) fixed-width ASCII files, read by their documented column positions; the NHIS 2017 Sample Adult and Family CSV files; and the NHIS 2019 public-use Linked Mortality Files (NHIS_[year]_MORT_2019_PUBLIC.dat), read by the layout in the NCHS codebook. NHIS survey files and documentation are available from the NCHS gateway at https://www.cdc.gov/nchs/nhis/documentation/index.html (1997–2018 releases via the CDC Archive, https://archive.cdc.gov/www_cdc_gov/nchs/nhis/1997-2018.htm); the public-use Linked Mortality Files and documentation are available at https://www.cdc.gov/nchs/data-linkage/mortality-public.htm. No other data inputs are used.

To replicate: download the source files into <root>/analysis/nhis_raw/{2002, 2007, 2012, 2017}/ using the subfolder and file names that appear in program A1, and edit the root path defined at the top of each program. Two names reflect local-download artifacts of the authors’ machine: the 2002 and 2017 mortality files carry a “ (1)” suffix, and the 2007 family file sits in a folder named “familyxx_download” — rename the official downloads to match, or edit those paths. Software: R 4.6.0 with packages readr, dplyr, tidyr, survey, survival, and splines; random seeds are set inside the programs. Approximate run times: A1 ≈10 minutes; A2–A5 a few minutes each; A6 ≈3.5 hours (300 bootstrap replicates and 300 bias-analysis iterations); A7 ≈30 minutes. Program A3 is excerpted through the doubly robust model stage used in this article; the remainder of that source file contains exploratory analyses not reported here. Figures were rendered from the CSV outputs of these programs.

**A1. run_nhis_pooled_meditation_mortality.R.** Builds the pooled analytic files from the federal source files (fixed-width NHIS ASCII files, 2017 CSV files, and public-use Linked Mortality Files); harmonizes the meditation exposure and covariates across waves; fits the covariate-adjusted survey Cox models (binary and three-level exposures).

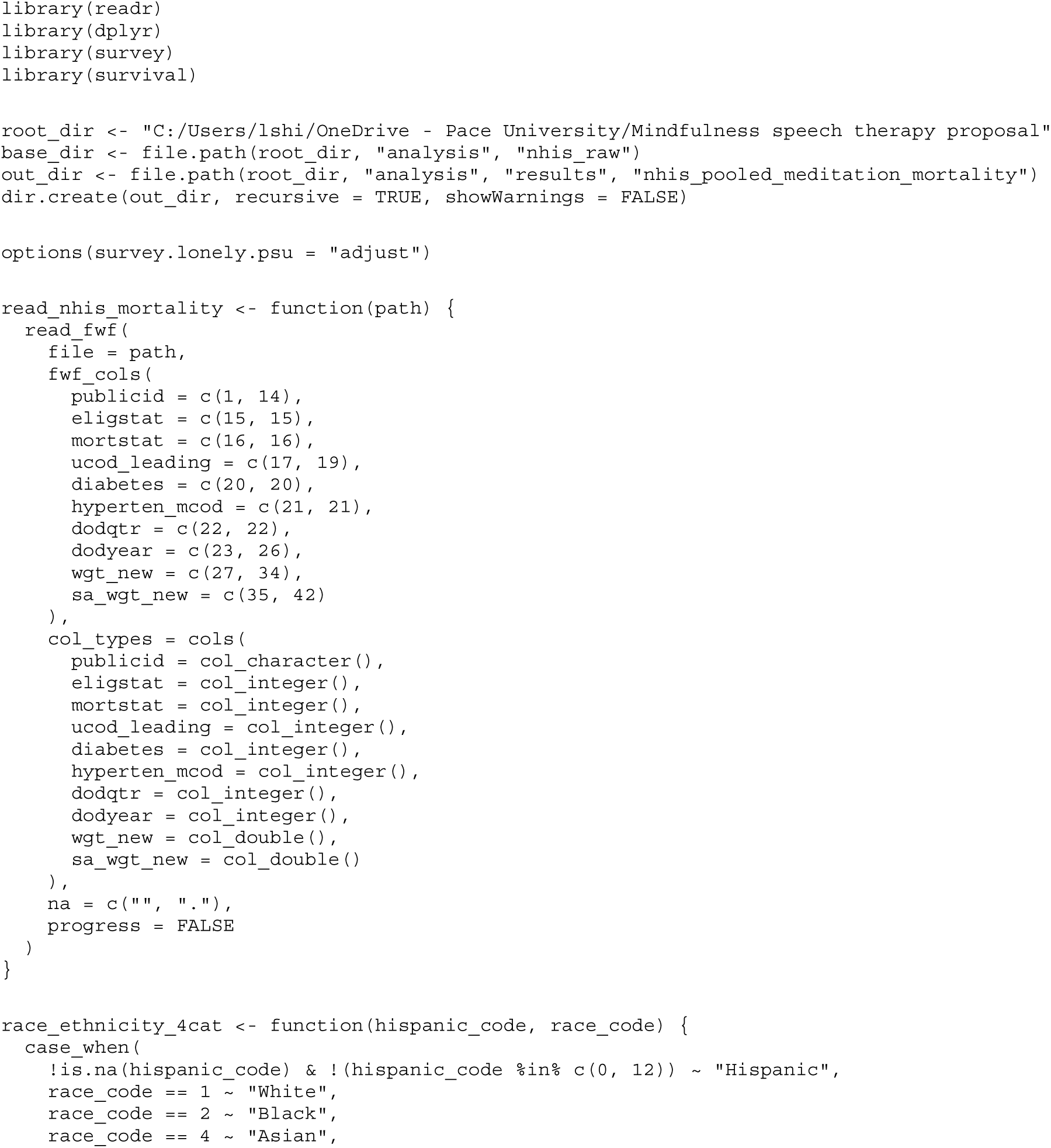
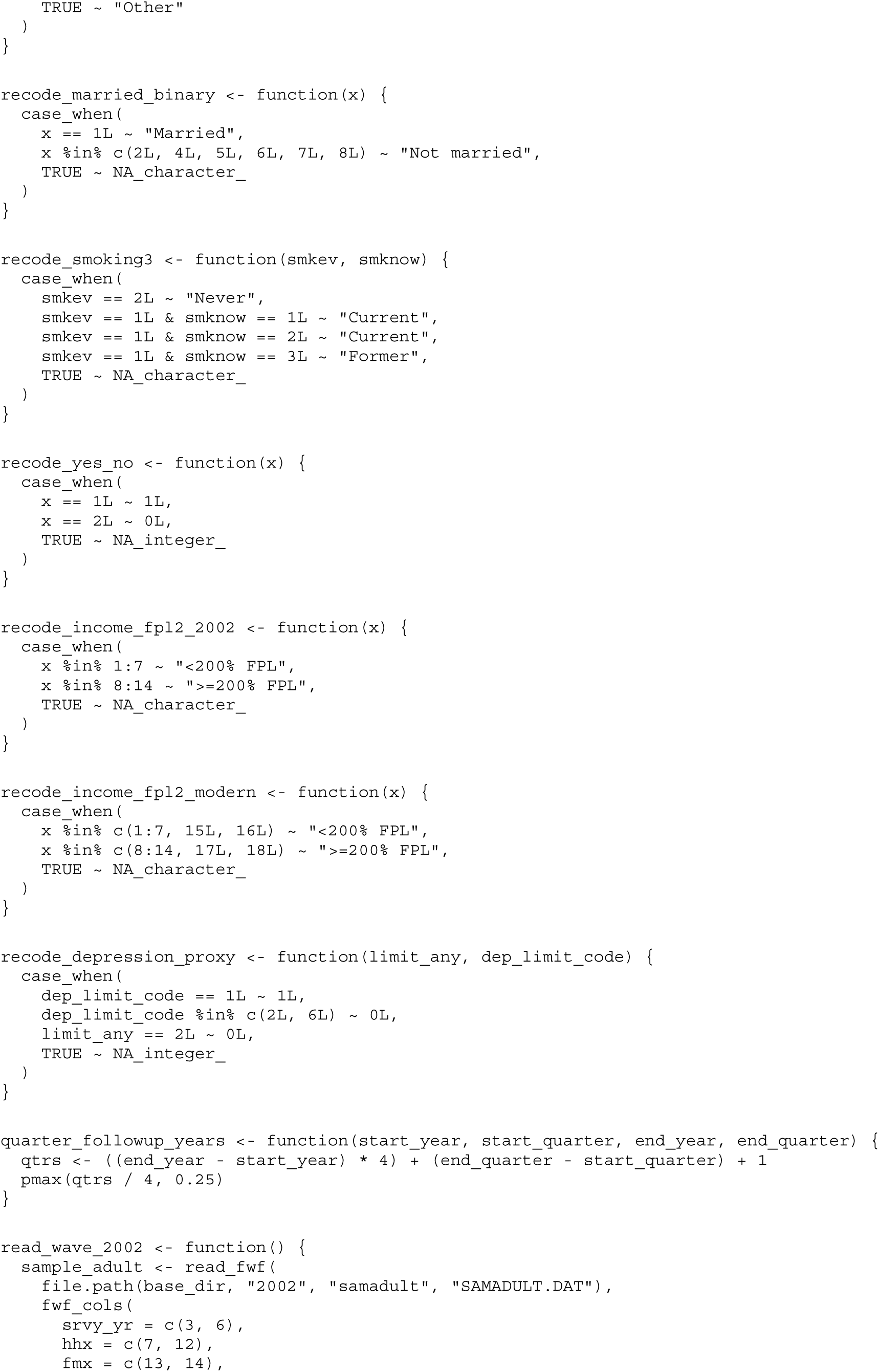
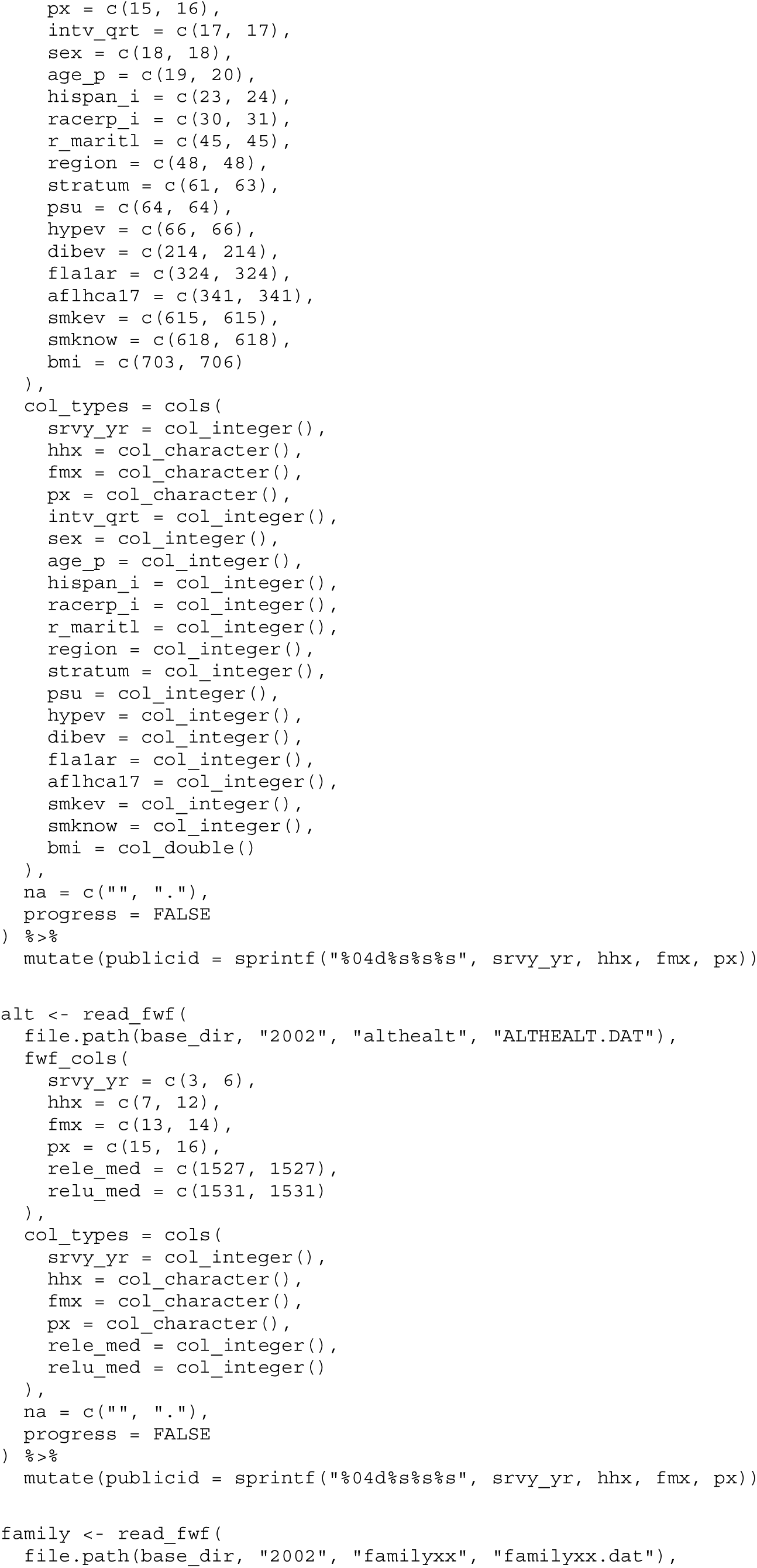
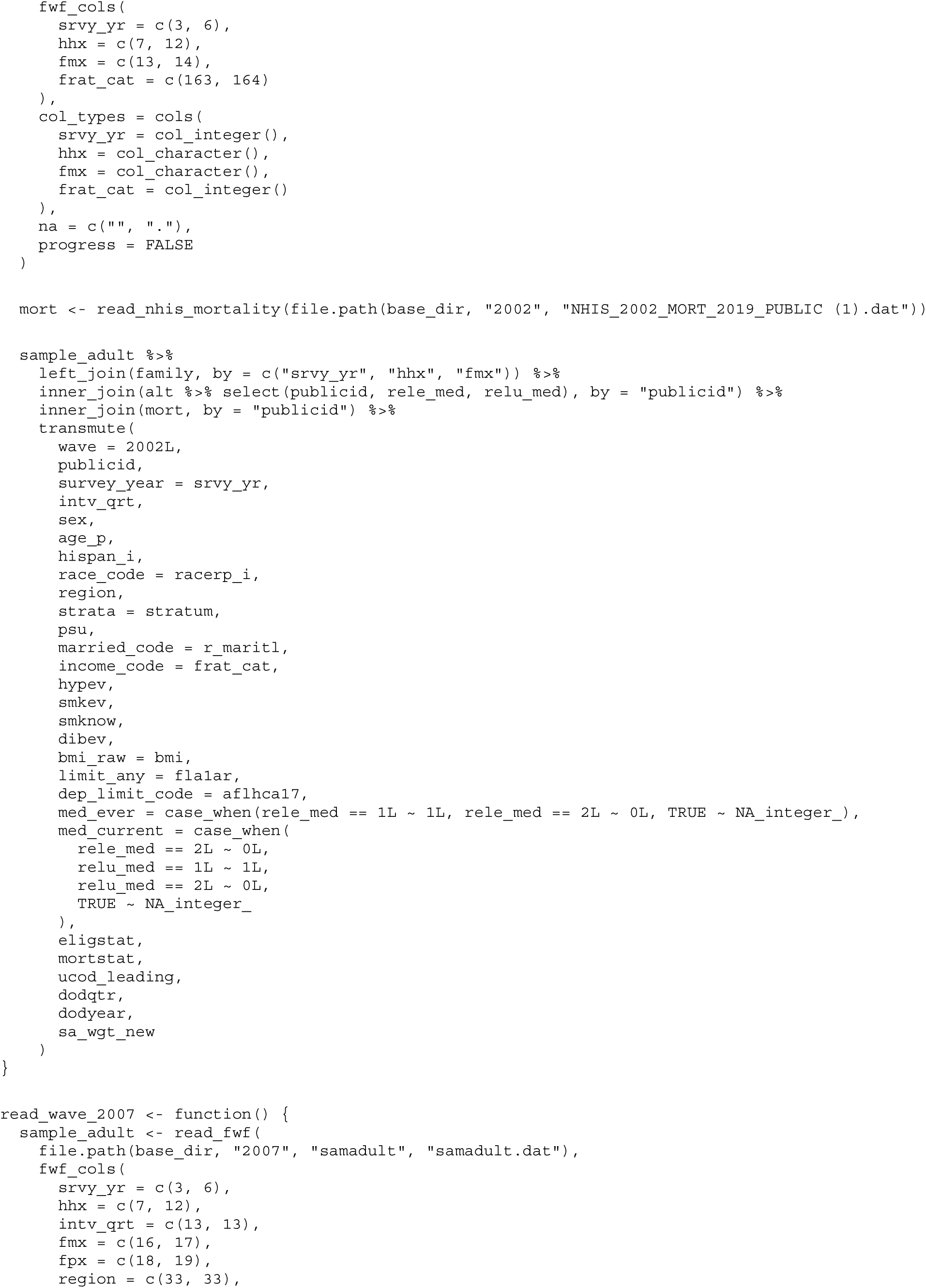
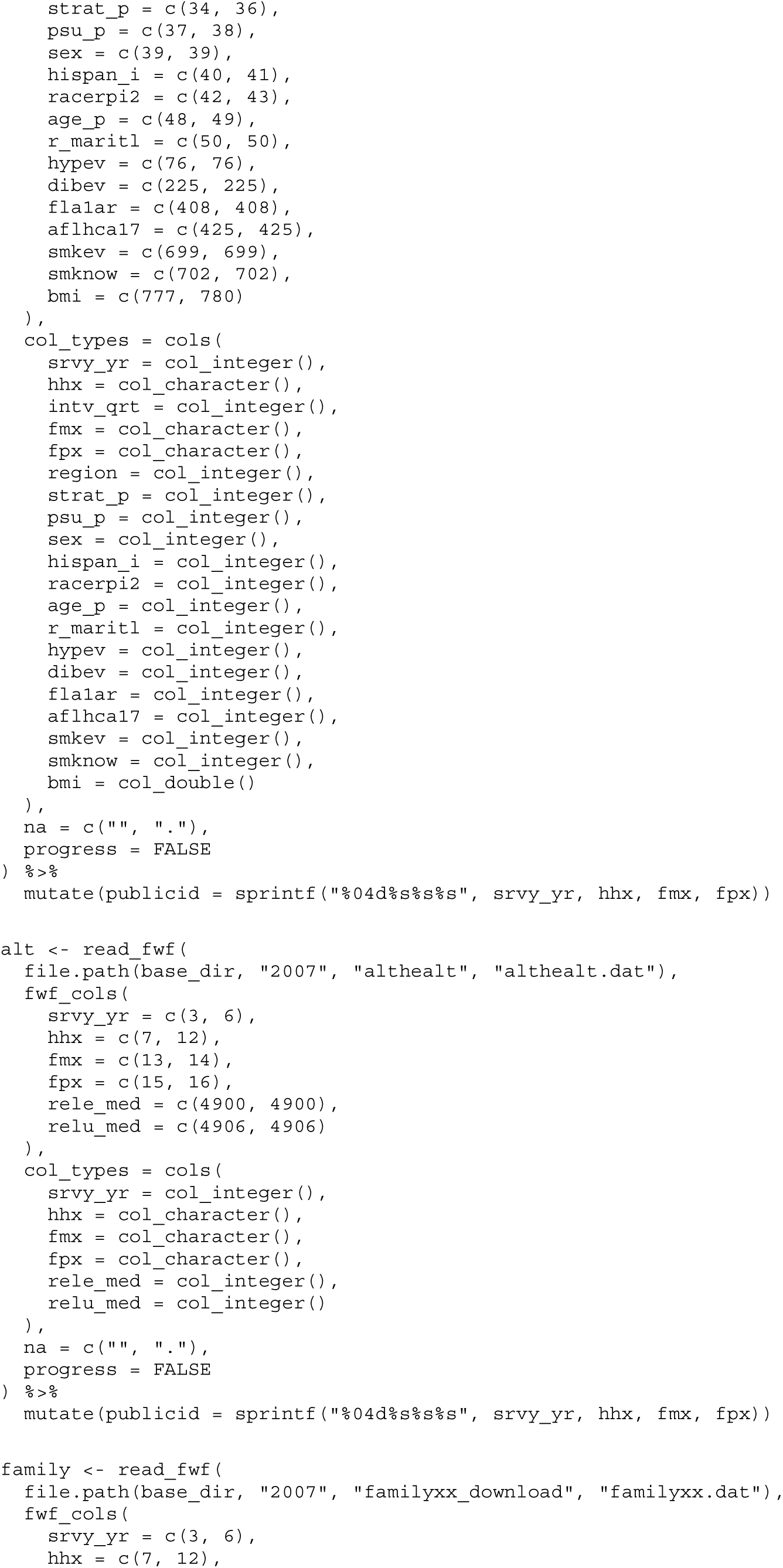
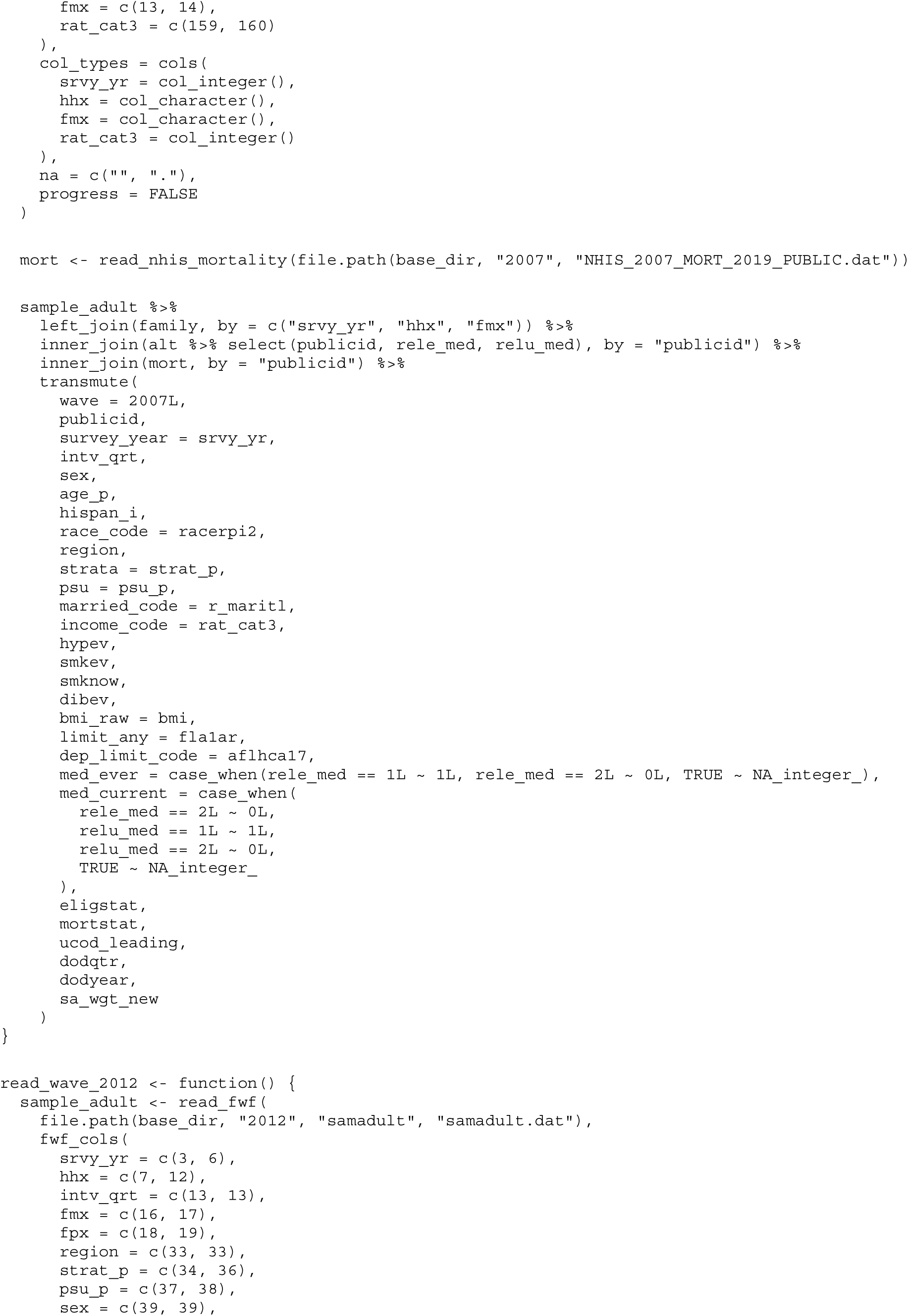
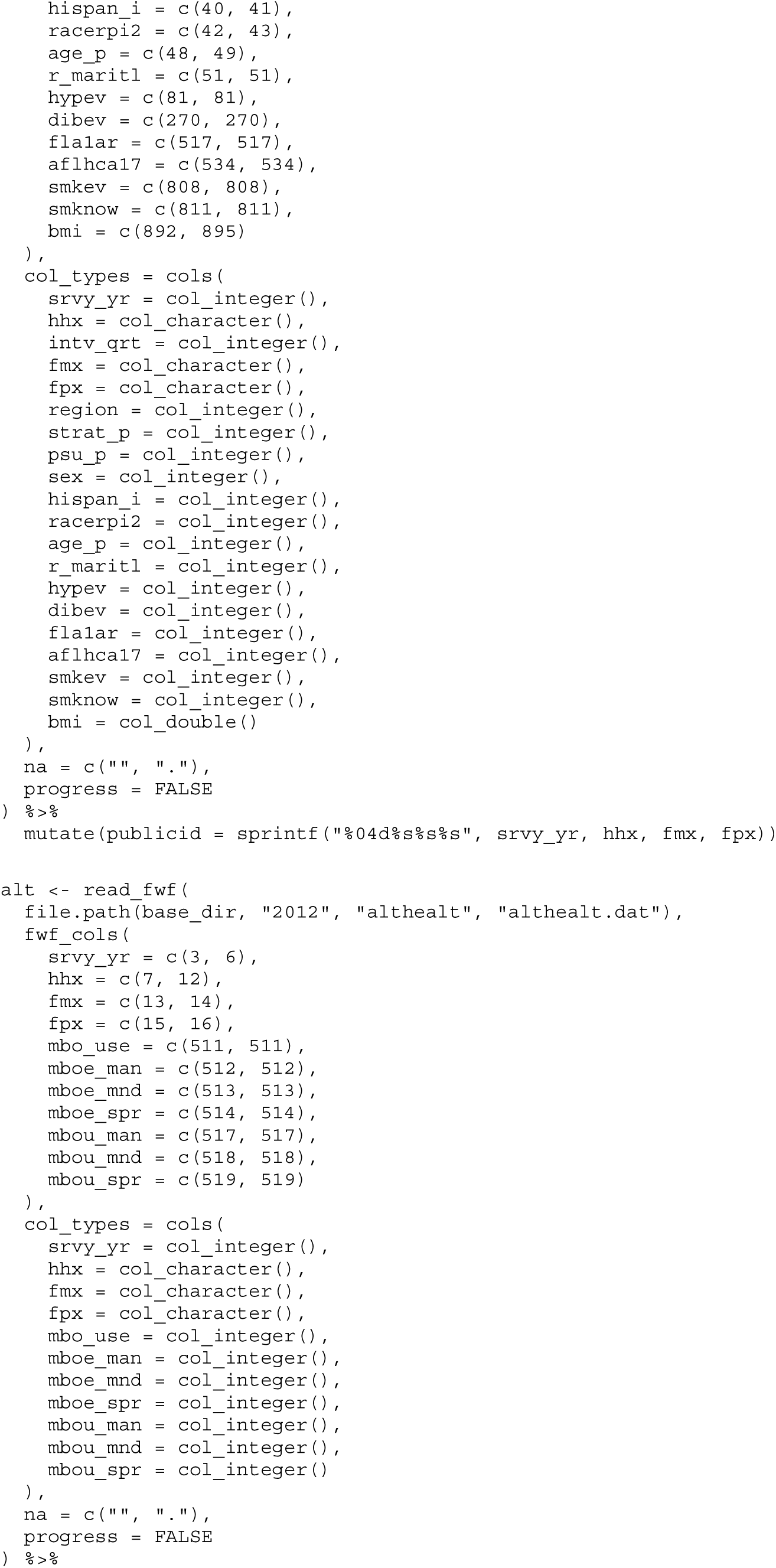
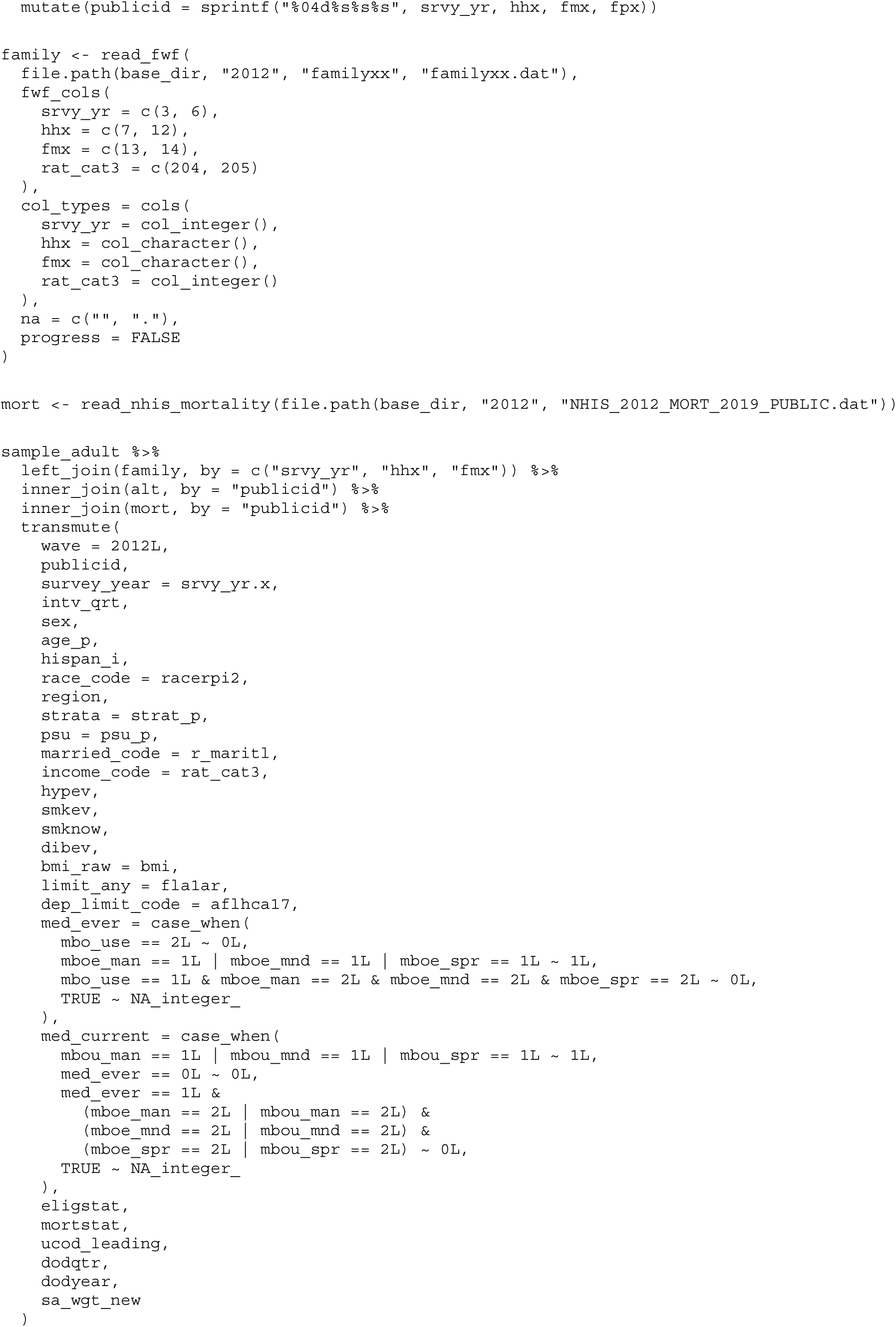
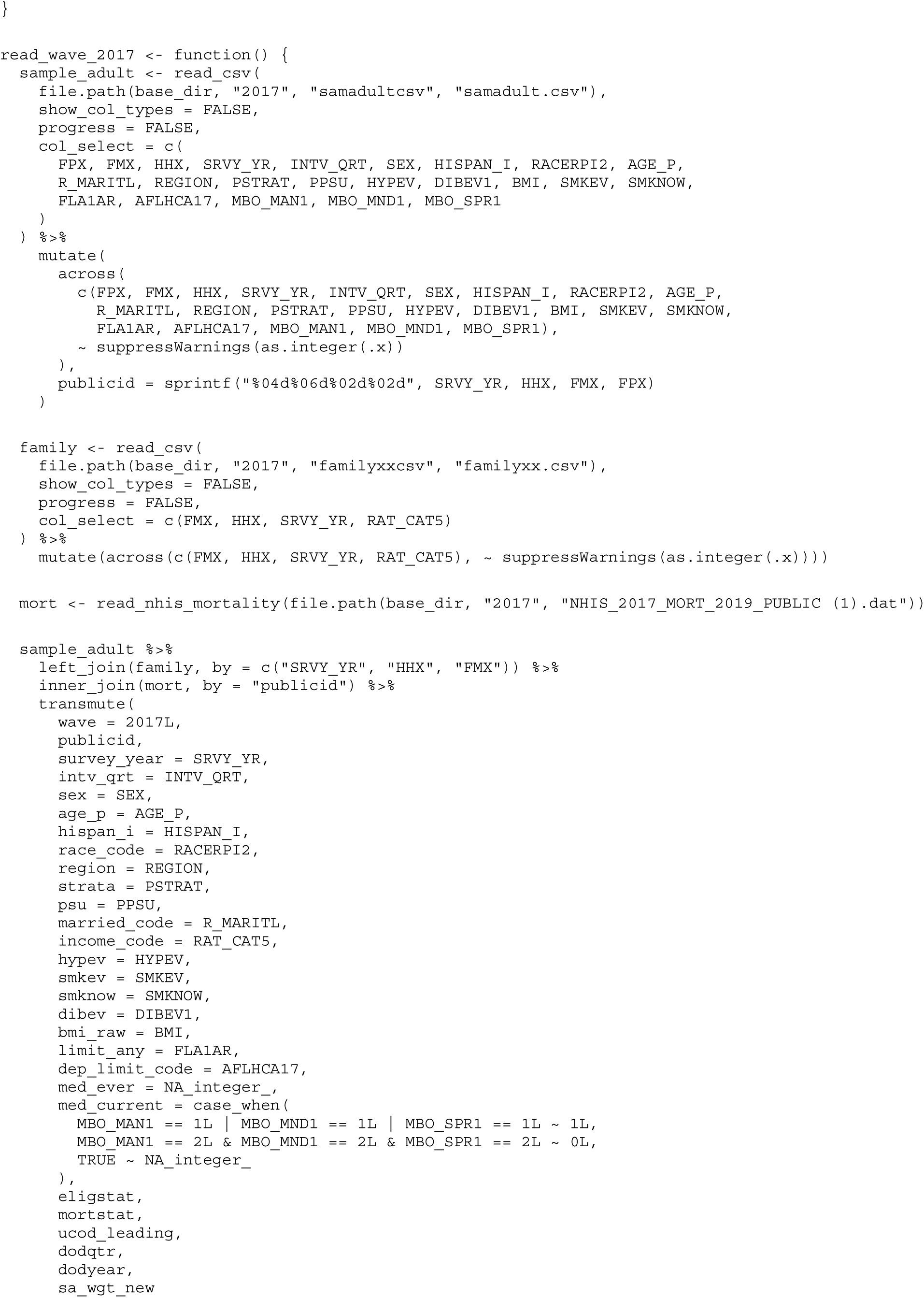
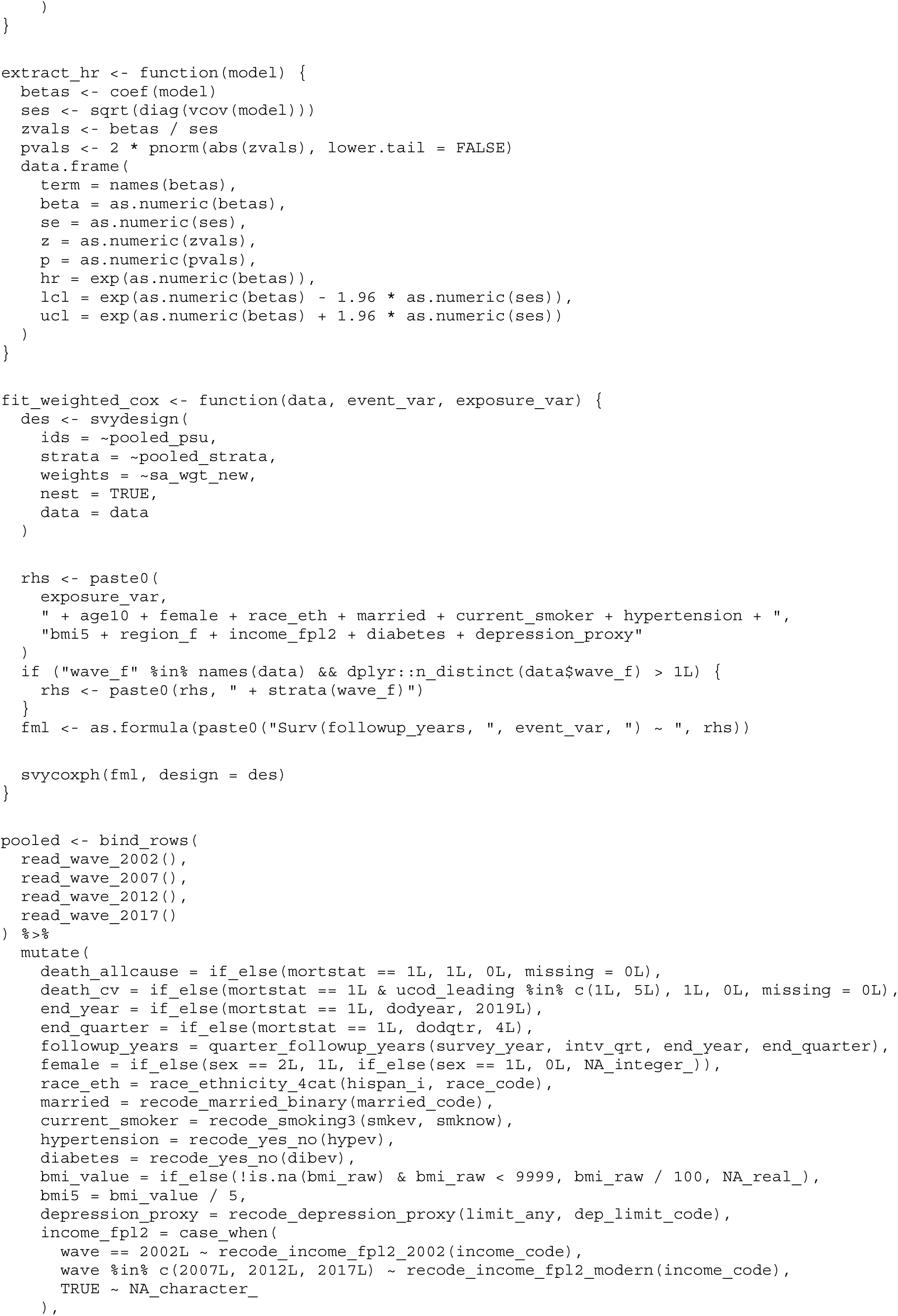
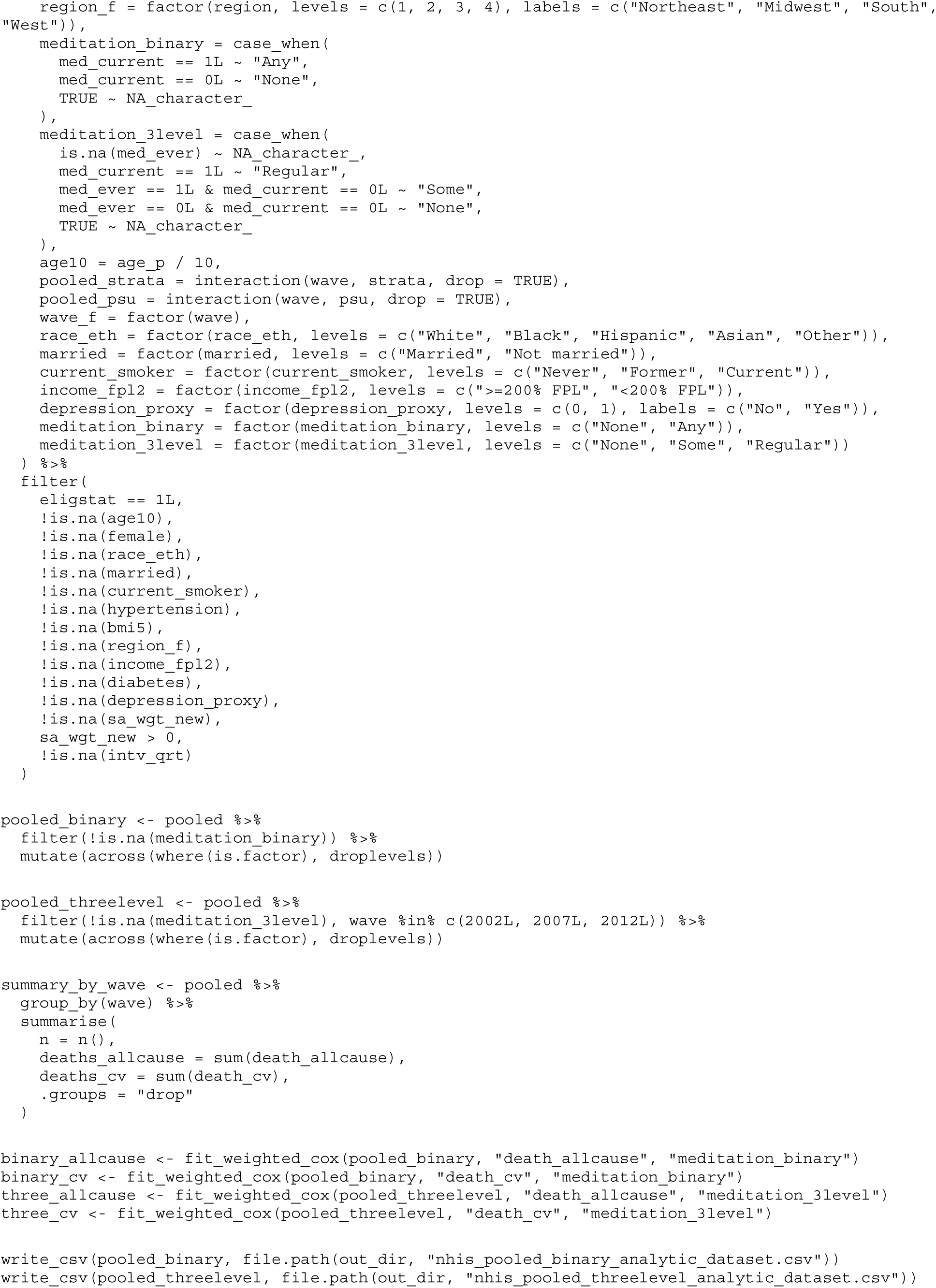
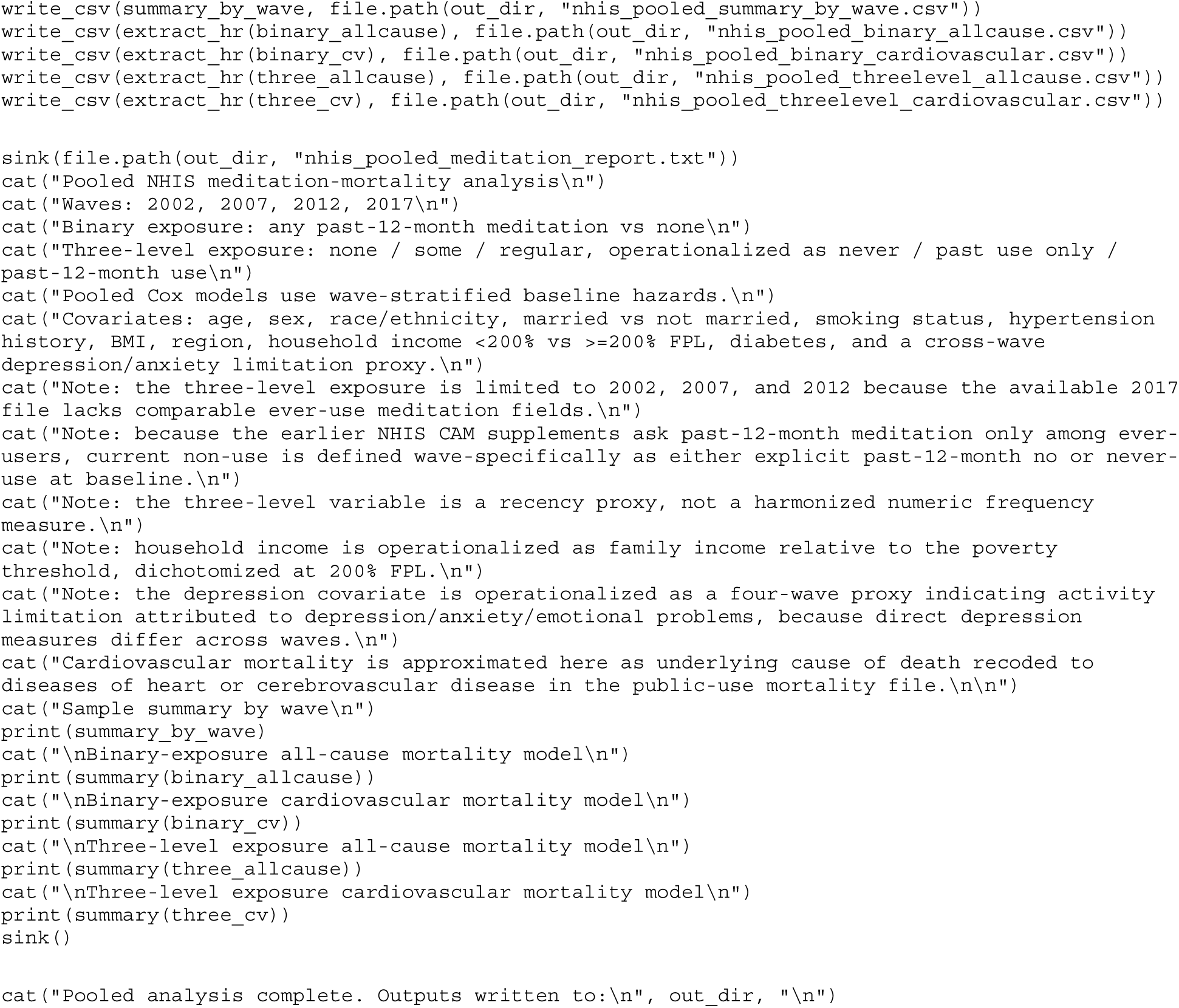

**A2. build_black55_hypertensive_teaching_dataset.R.** Constructs the subgroup analytic file (non-Hispanic Black adults aged ≥*55 years with hypertension) from the pooled analytic file, appending two auxiliary lifestyle variables read from the same federal source files*.

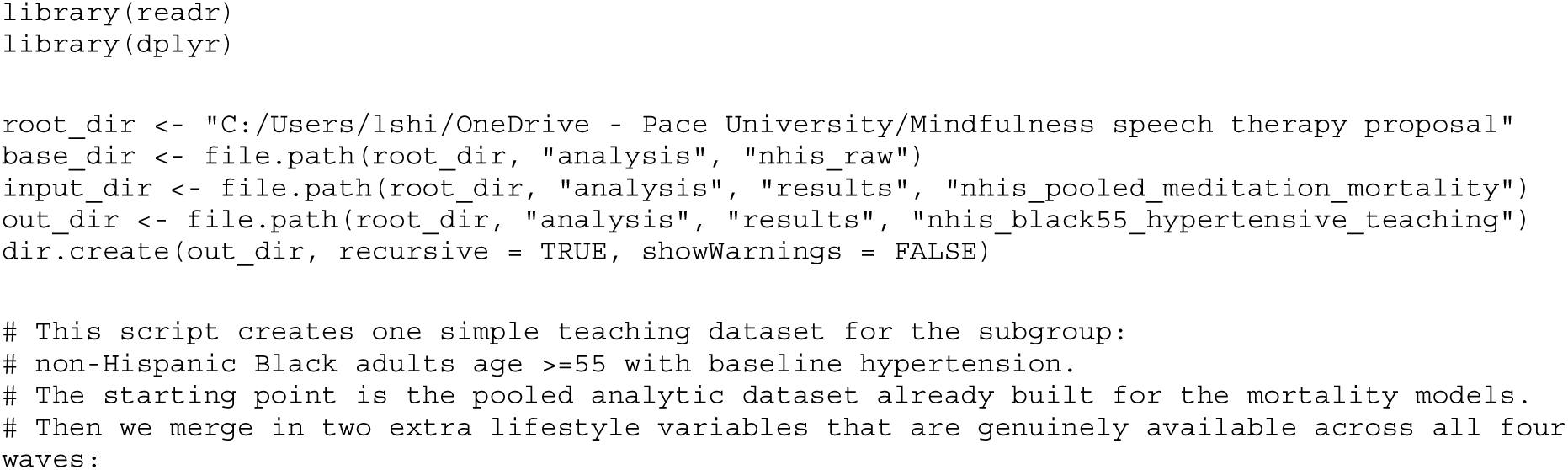
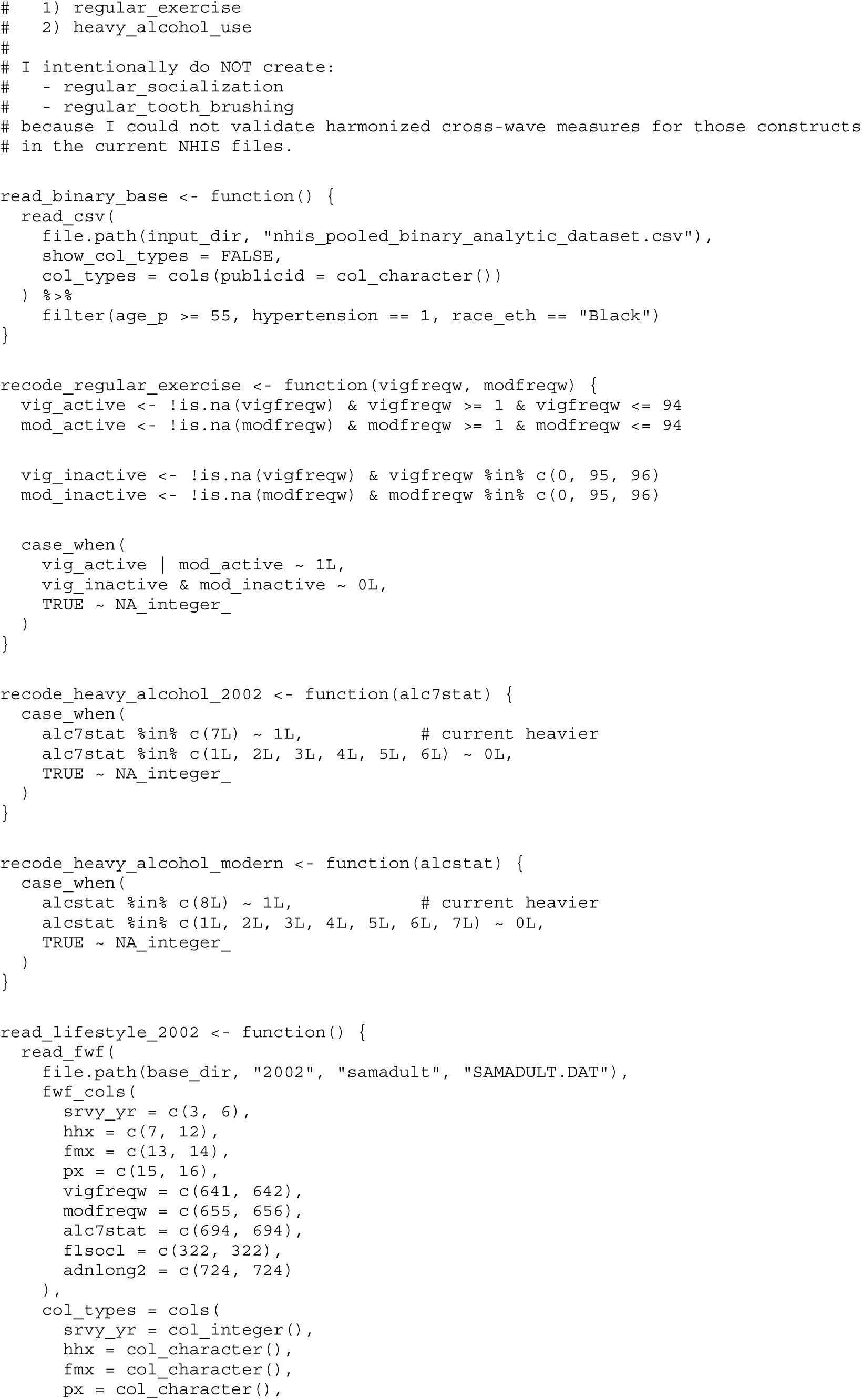
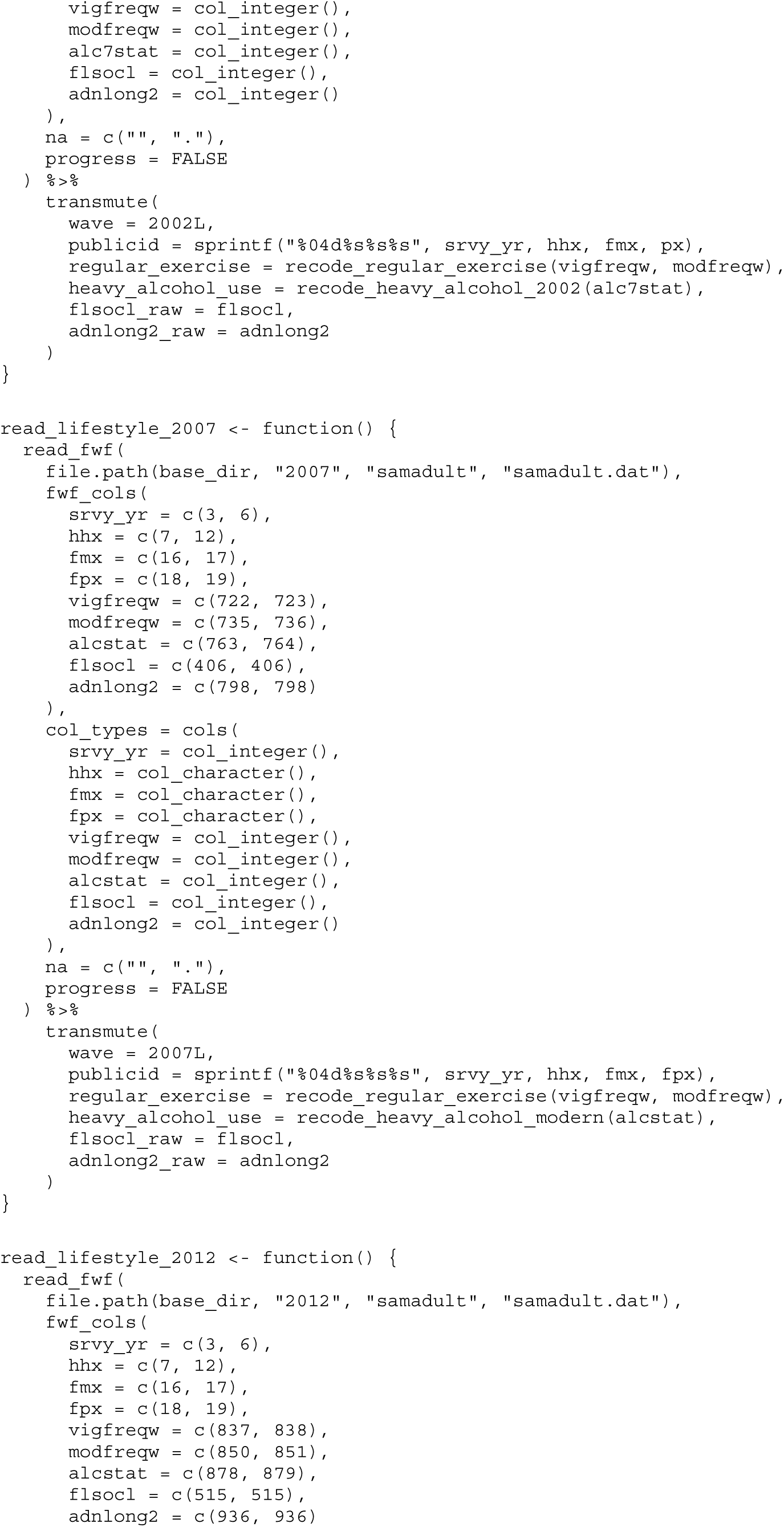
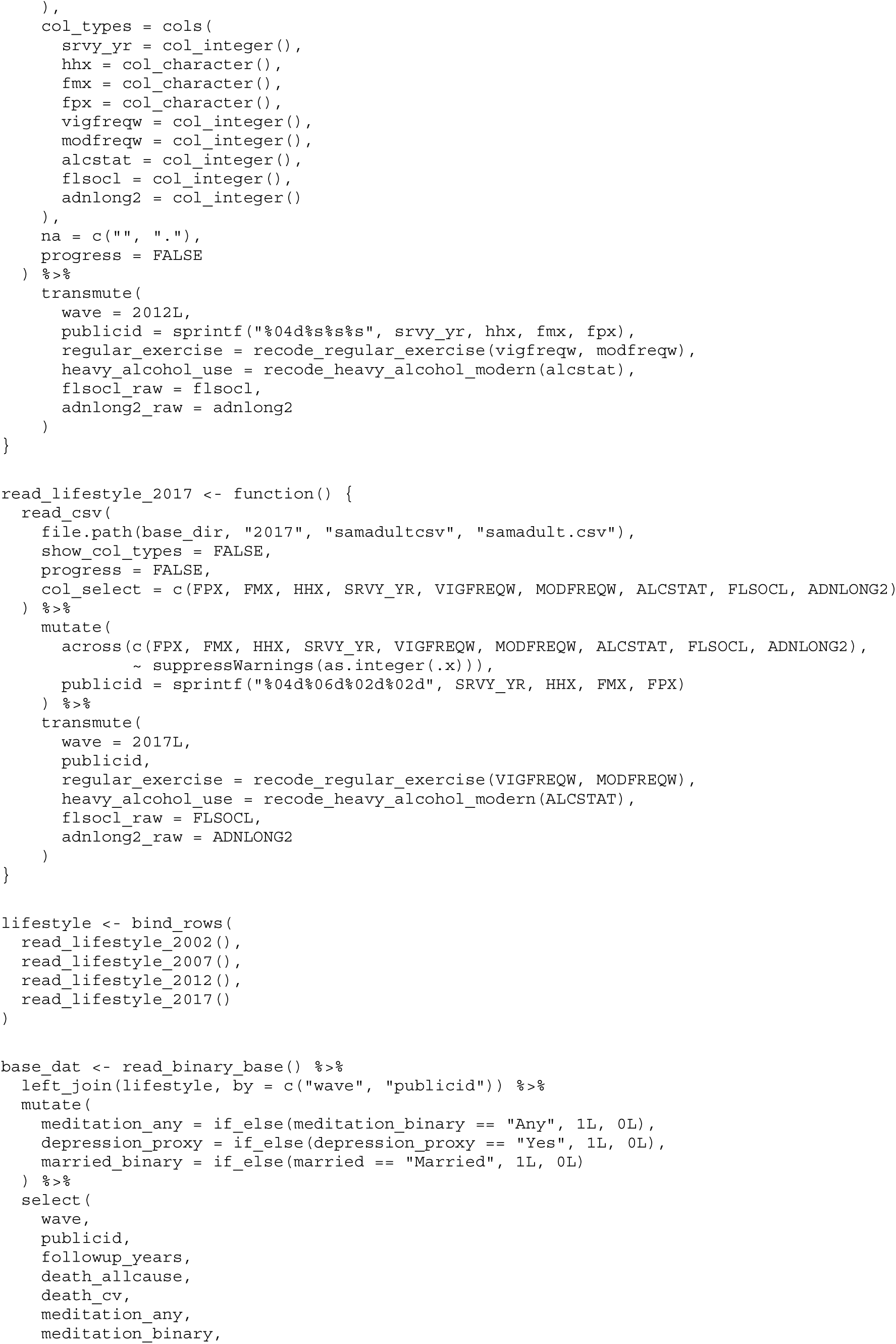
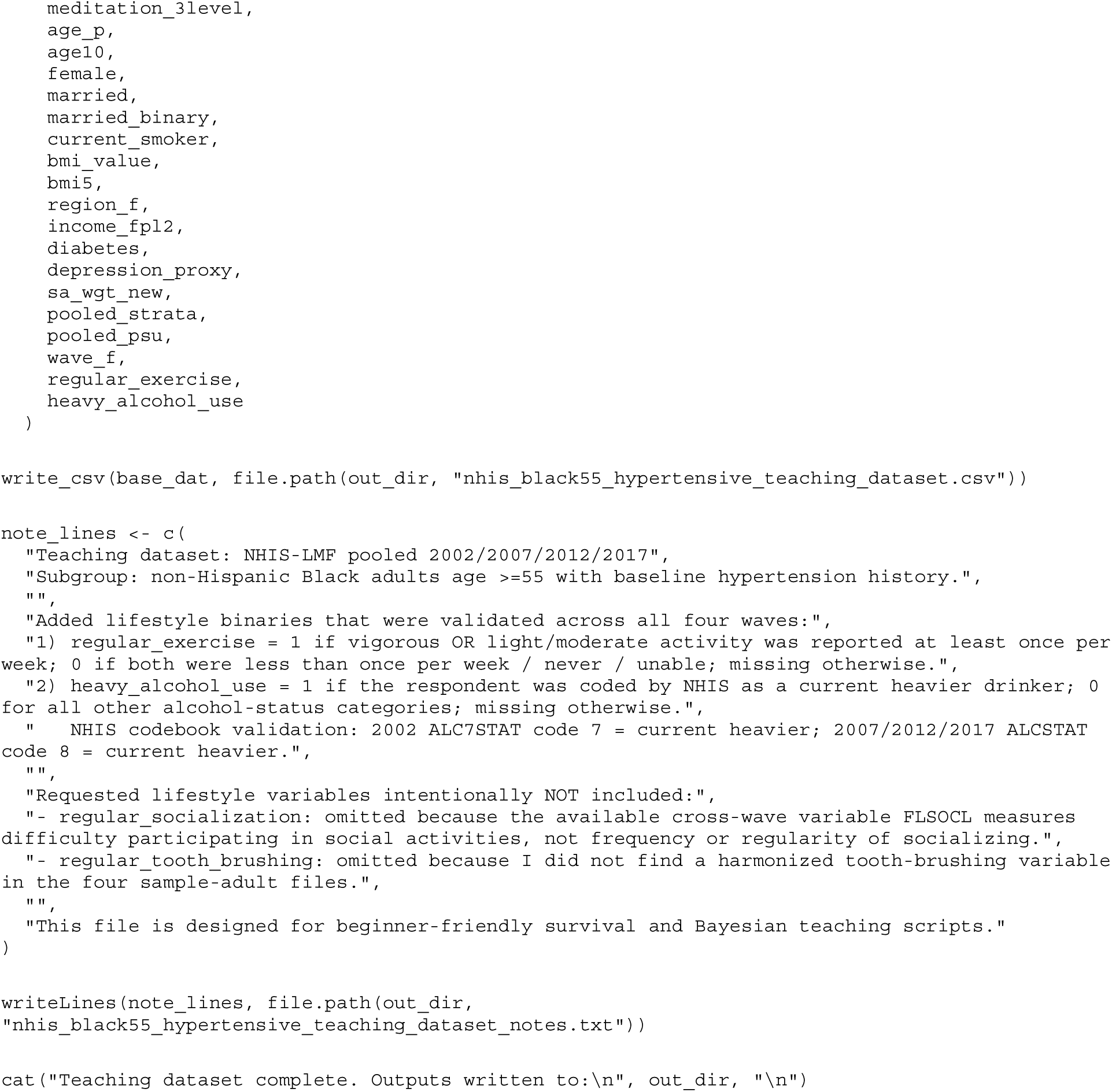

**A3. run_black55_dr_qba.R (excerpt).** Subgroup doubly robust survey Cox models whose estimates serve as the likelihood for the Bayesian synthesis; excerpted through the model stage used in this article.

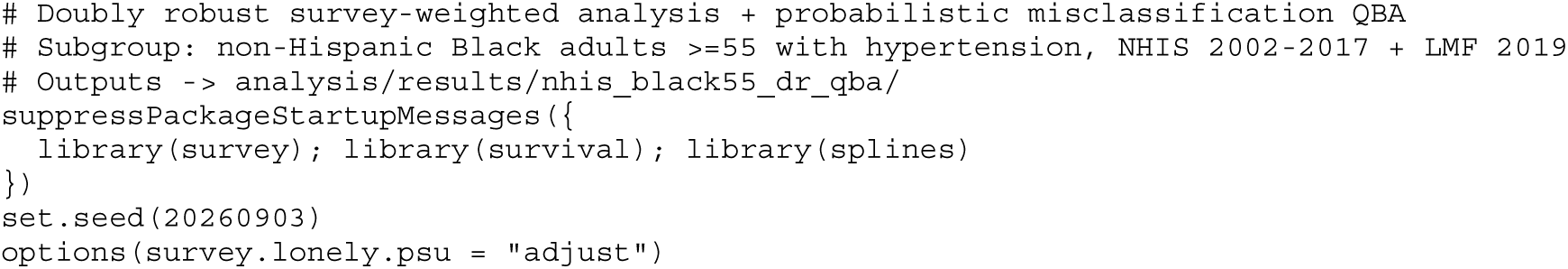
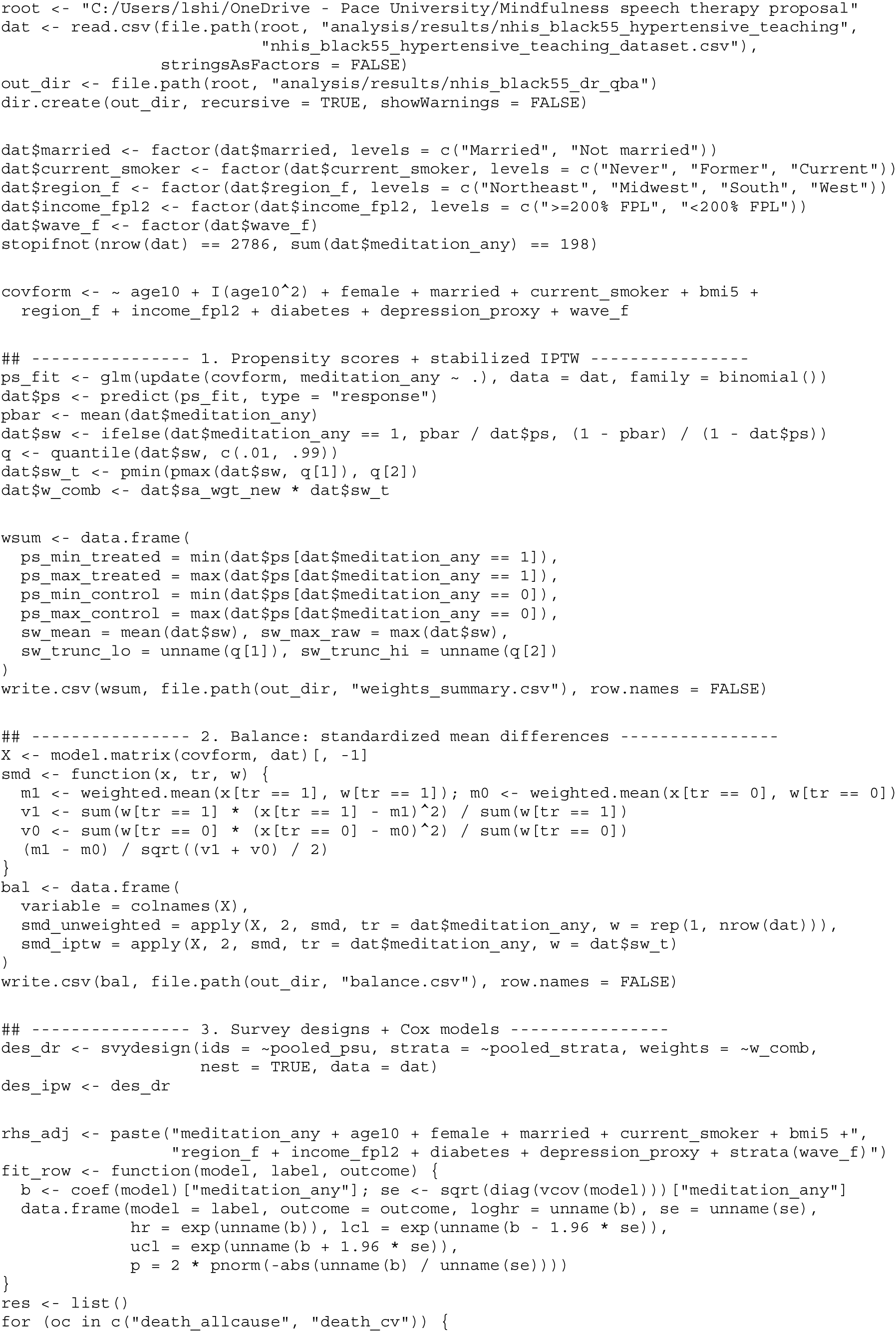
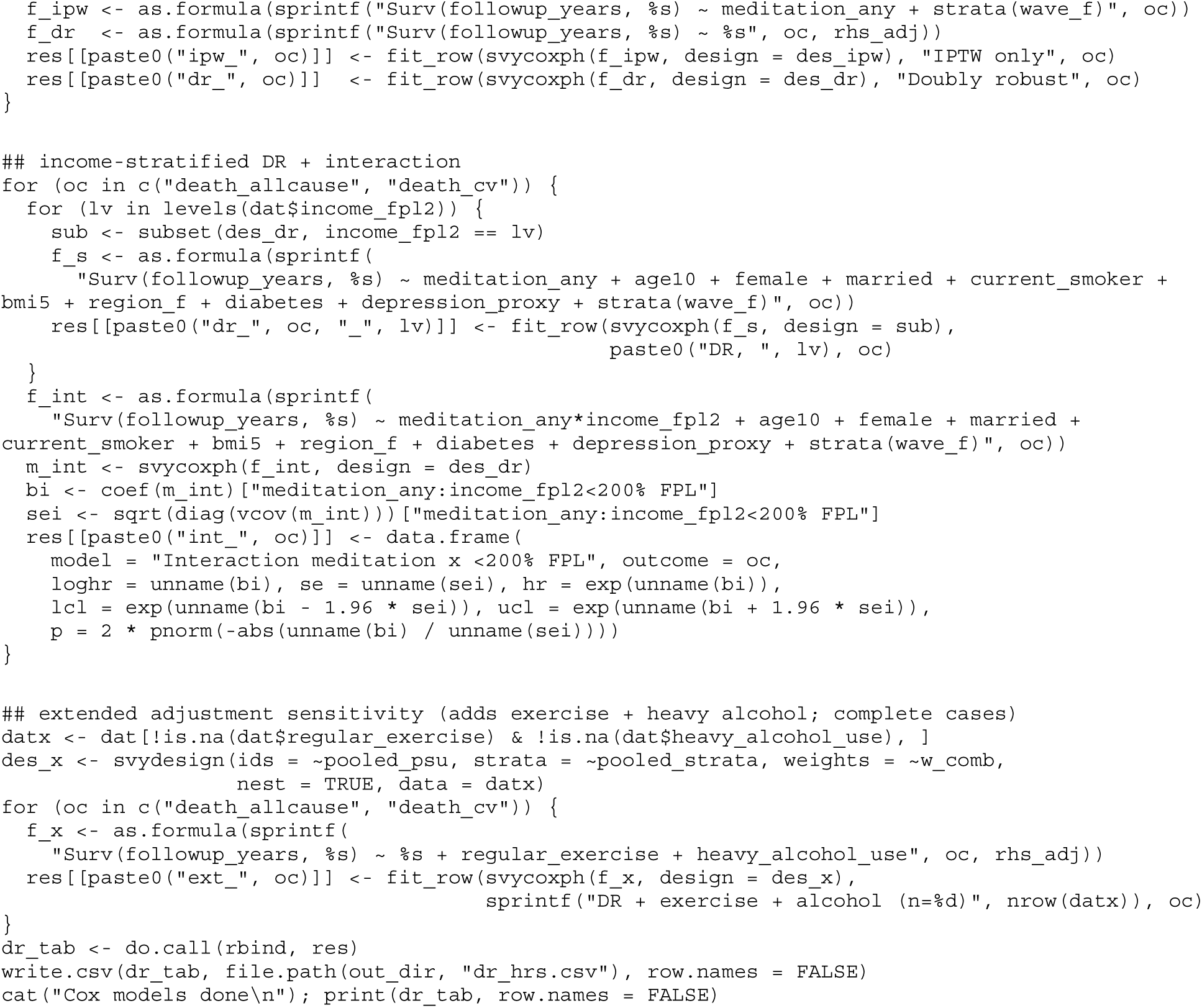

**A4. run_pooled_dr_core.R.** Full-sample analyses: propensity scores, stabilized weights, balance diagnostics, doubly robust and weighting-only models, income-stratified models and interaction, Kaplan–Meier estimates, E-values, and the trial-informed Bayesian synthesis for the subgroup.

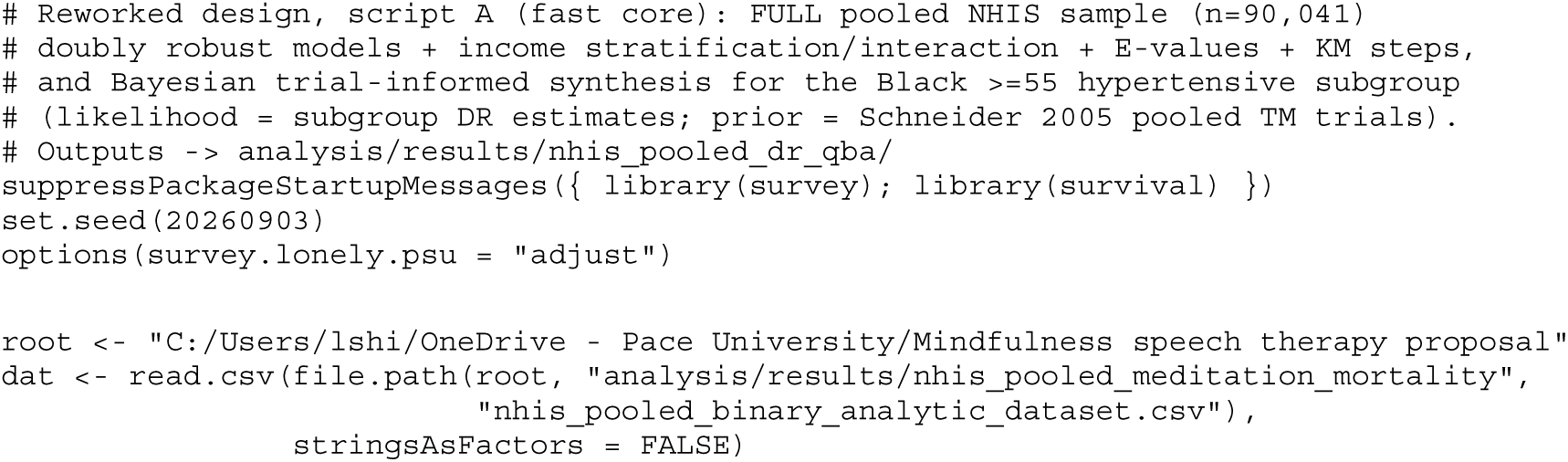
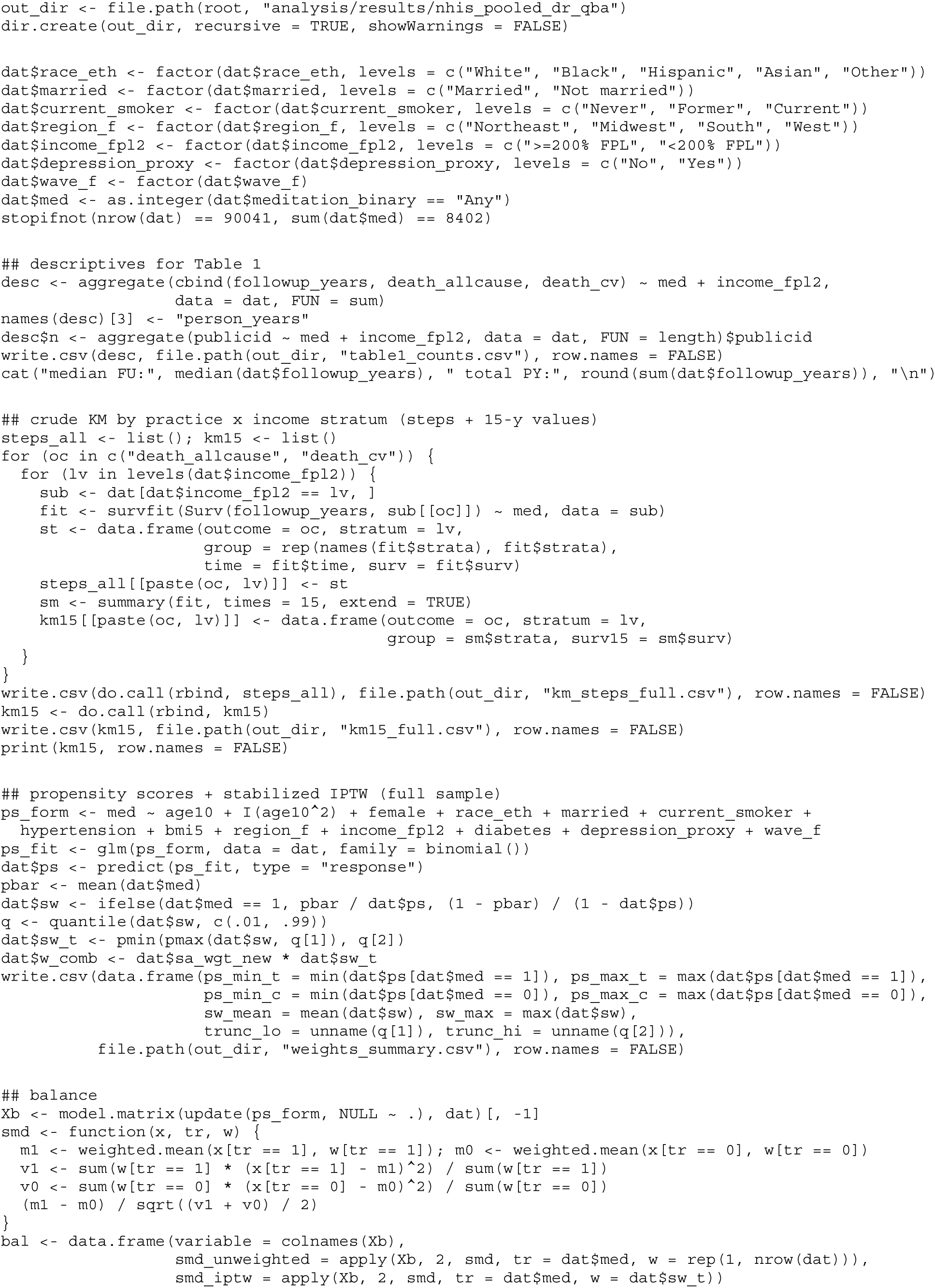
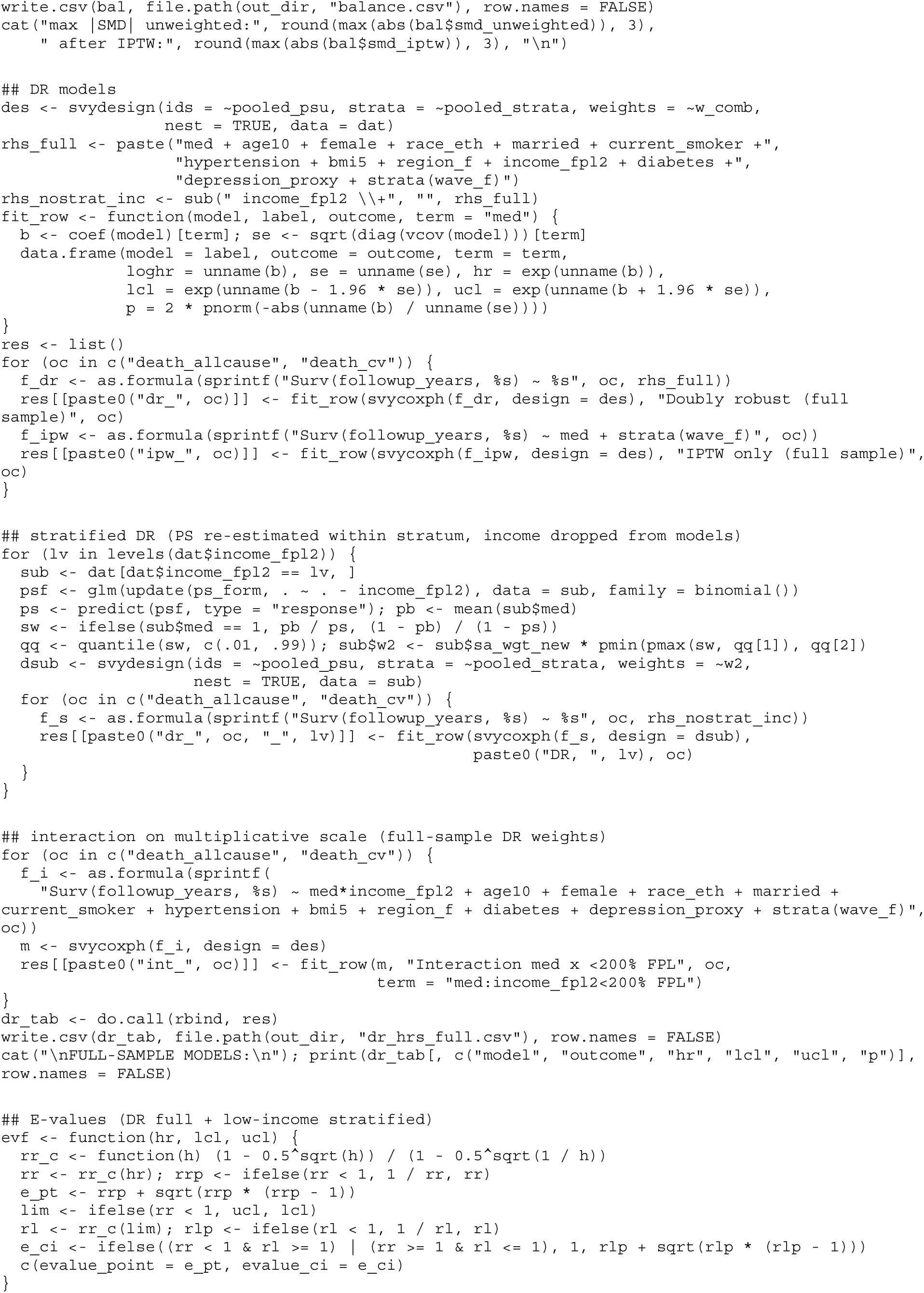
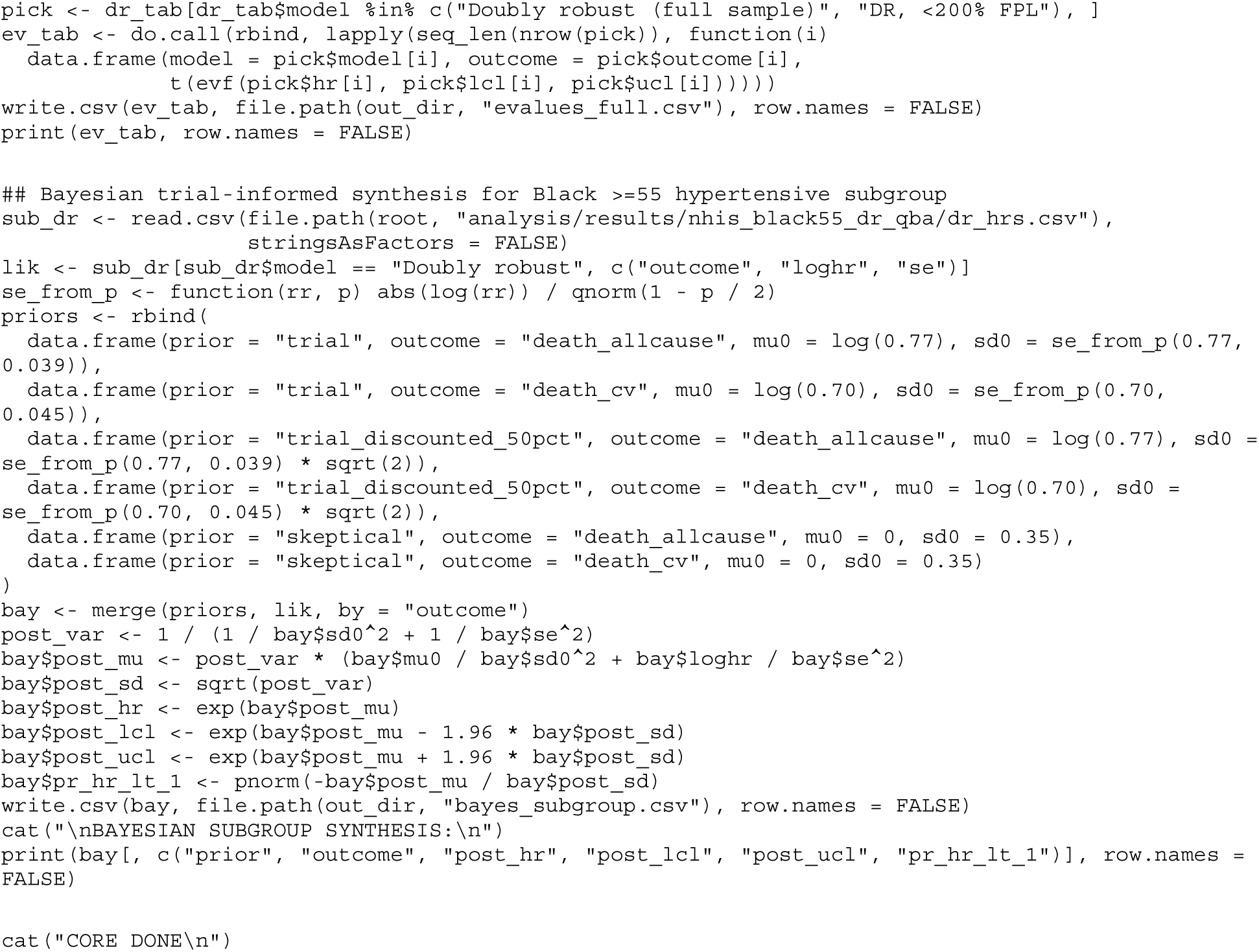

**A5. run_pooled_dr_minimal.R.** Minimal-adjustment (total-association) companion model conditioning only on pre-exposure demographic and socioeconomic covariates.

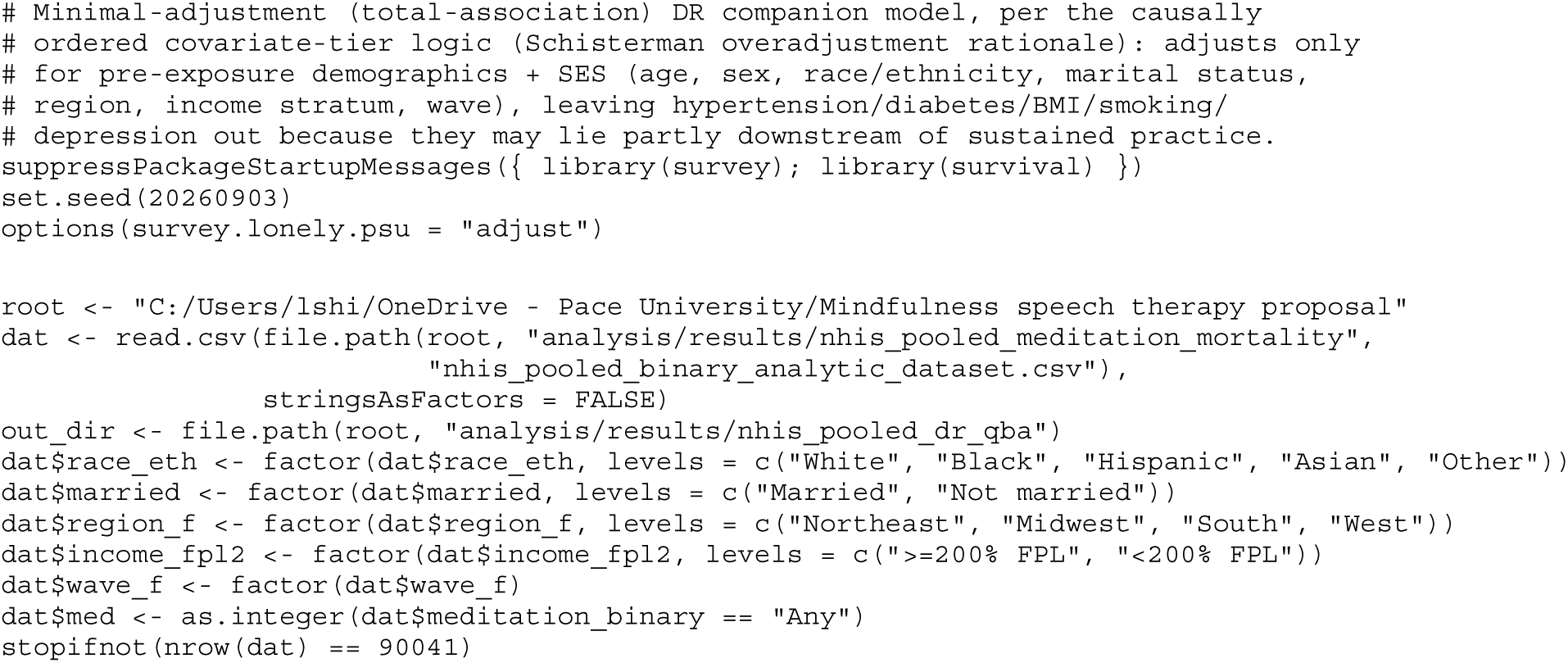
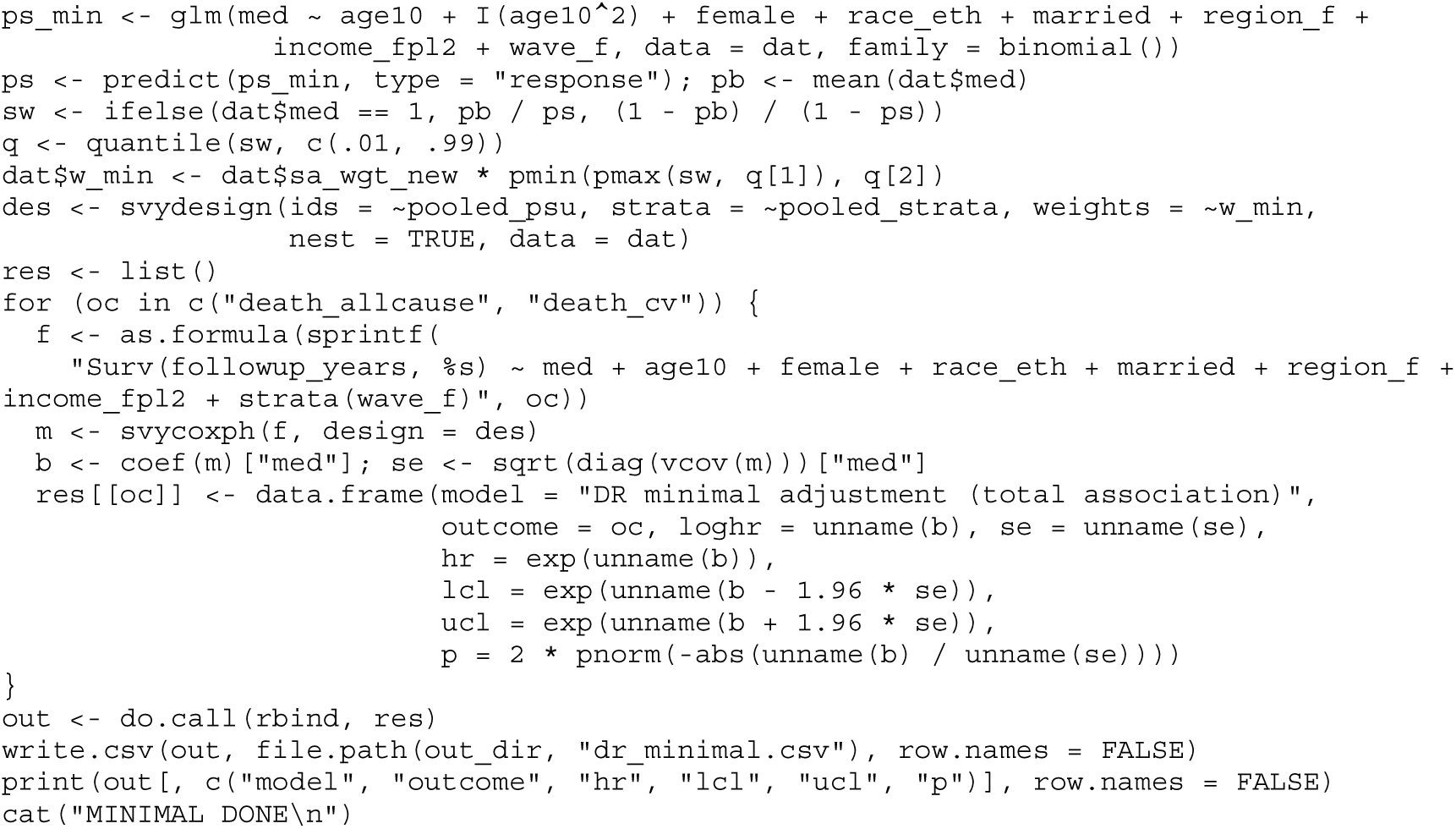

**A6. run_pooled_std_qba.R.** Standardized 15-year risks by g-computation with a cluster multiplier bootstrap, and the nondifferential probabilistic misclassification analysis.

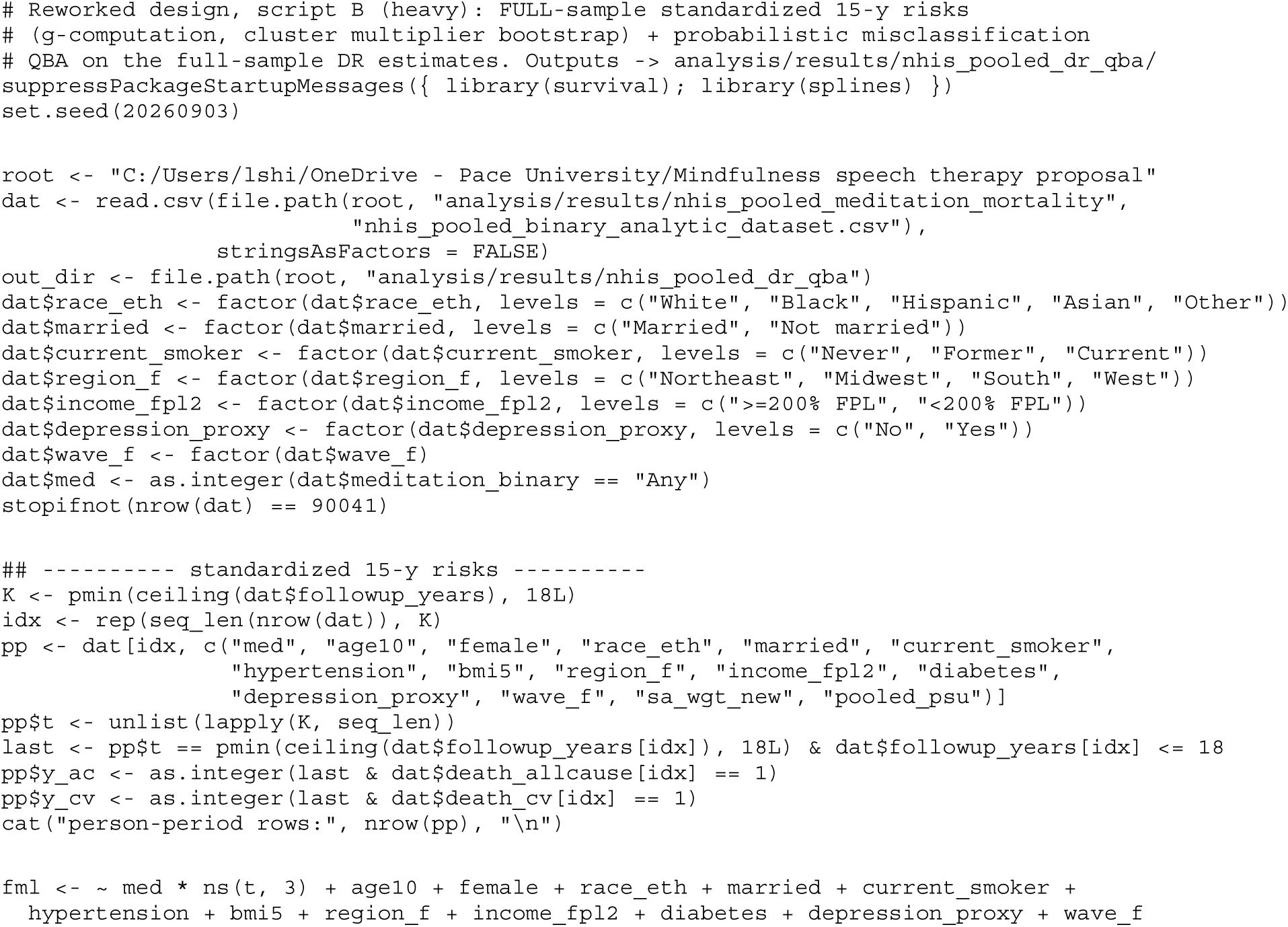
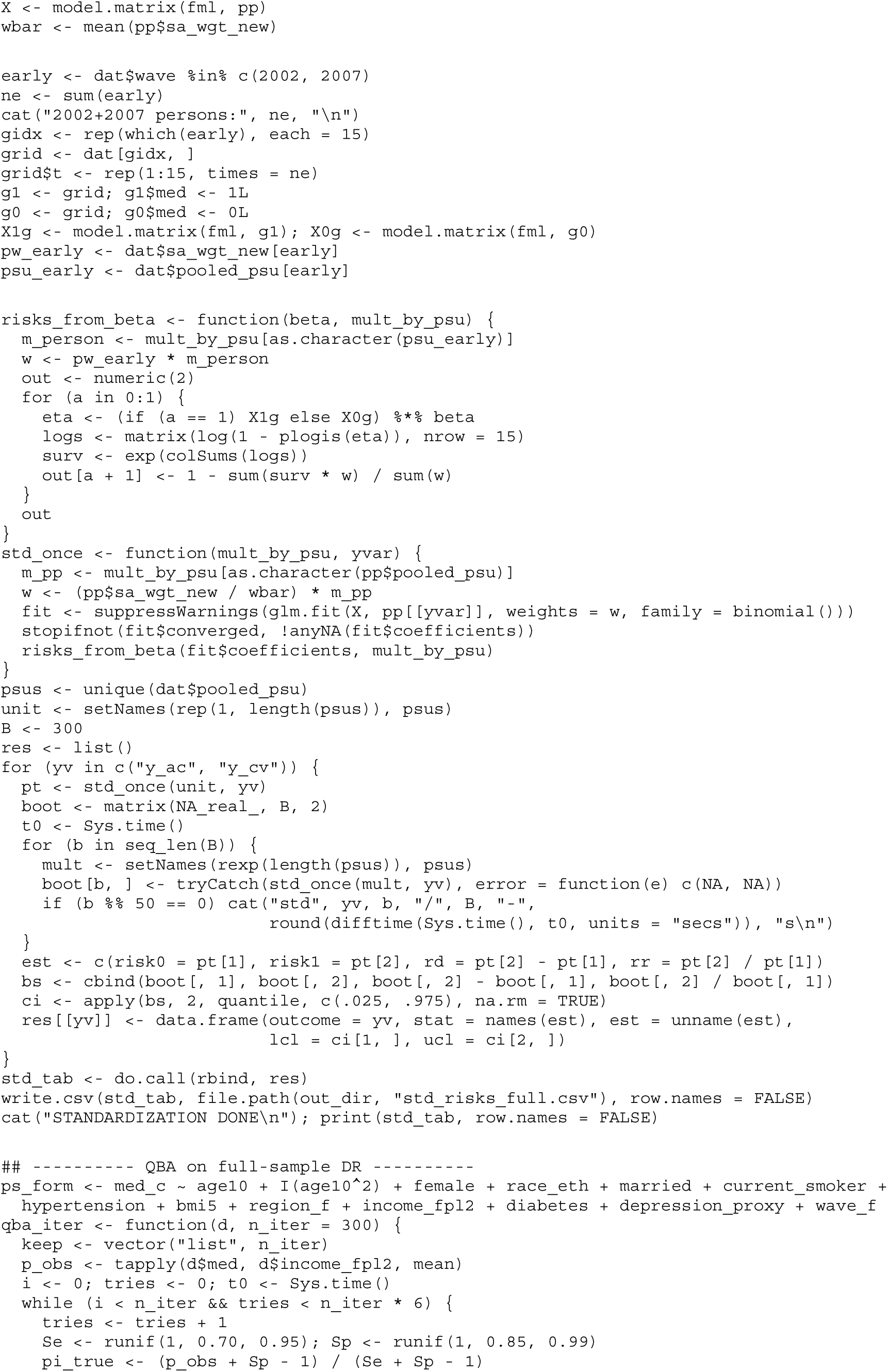
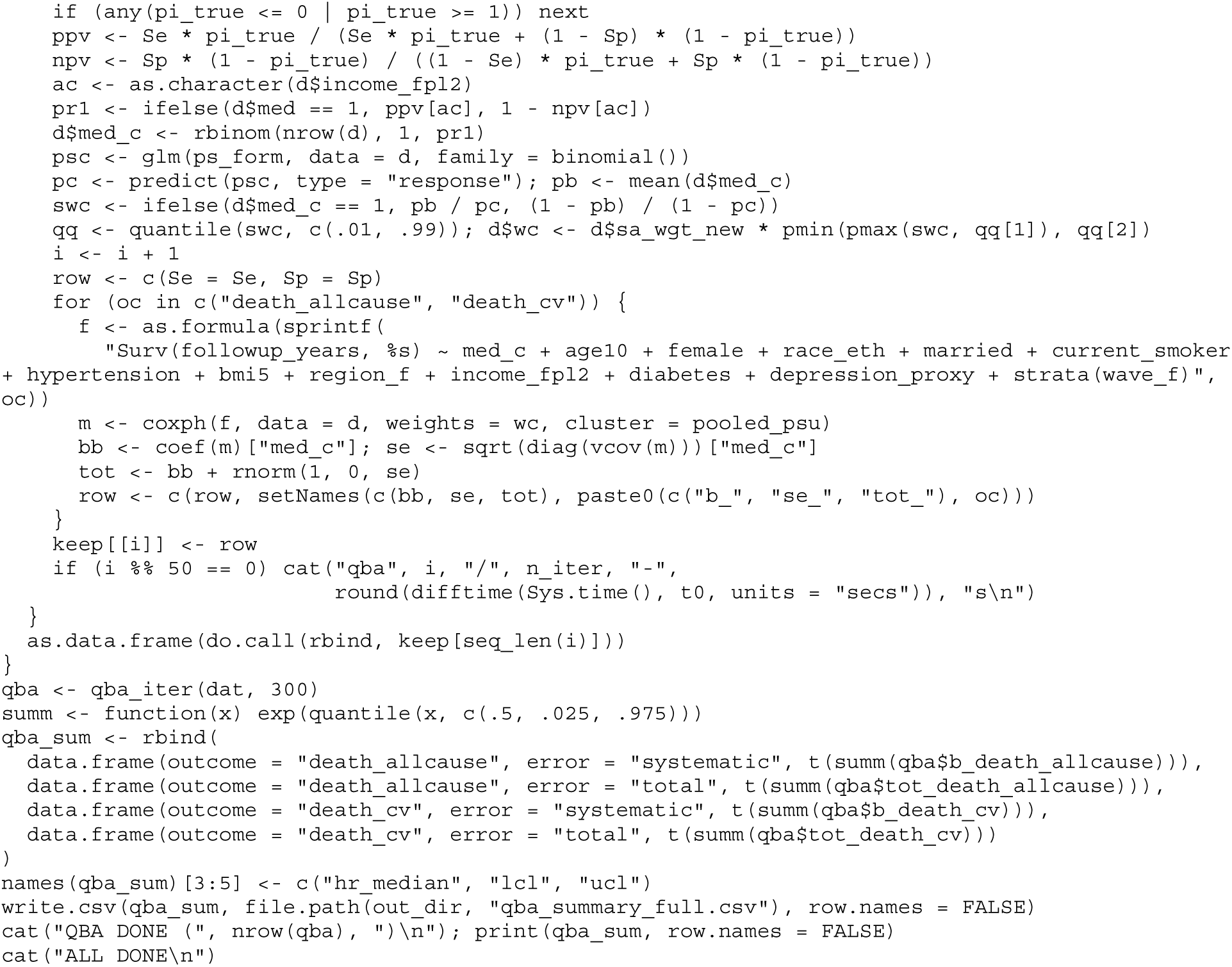

**A7. run_pooled_qba_differential.R.** Differential-by-income misclassification scenario targeting the practice-by-income interaction estimate.

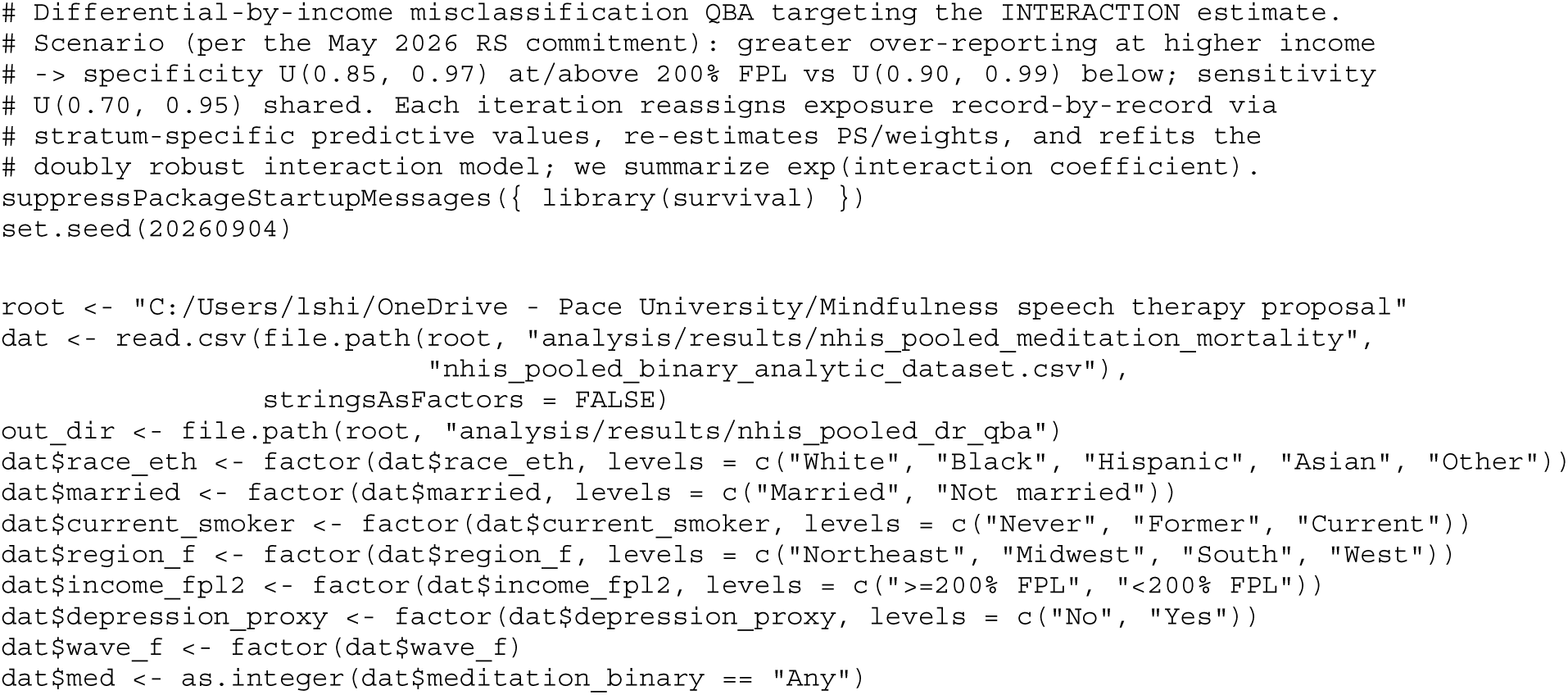
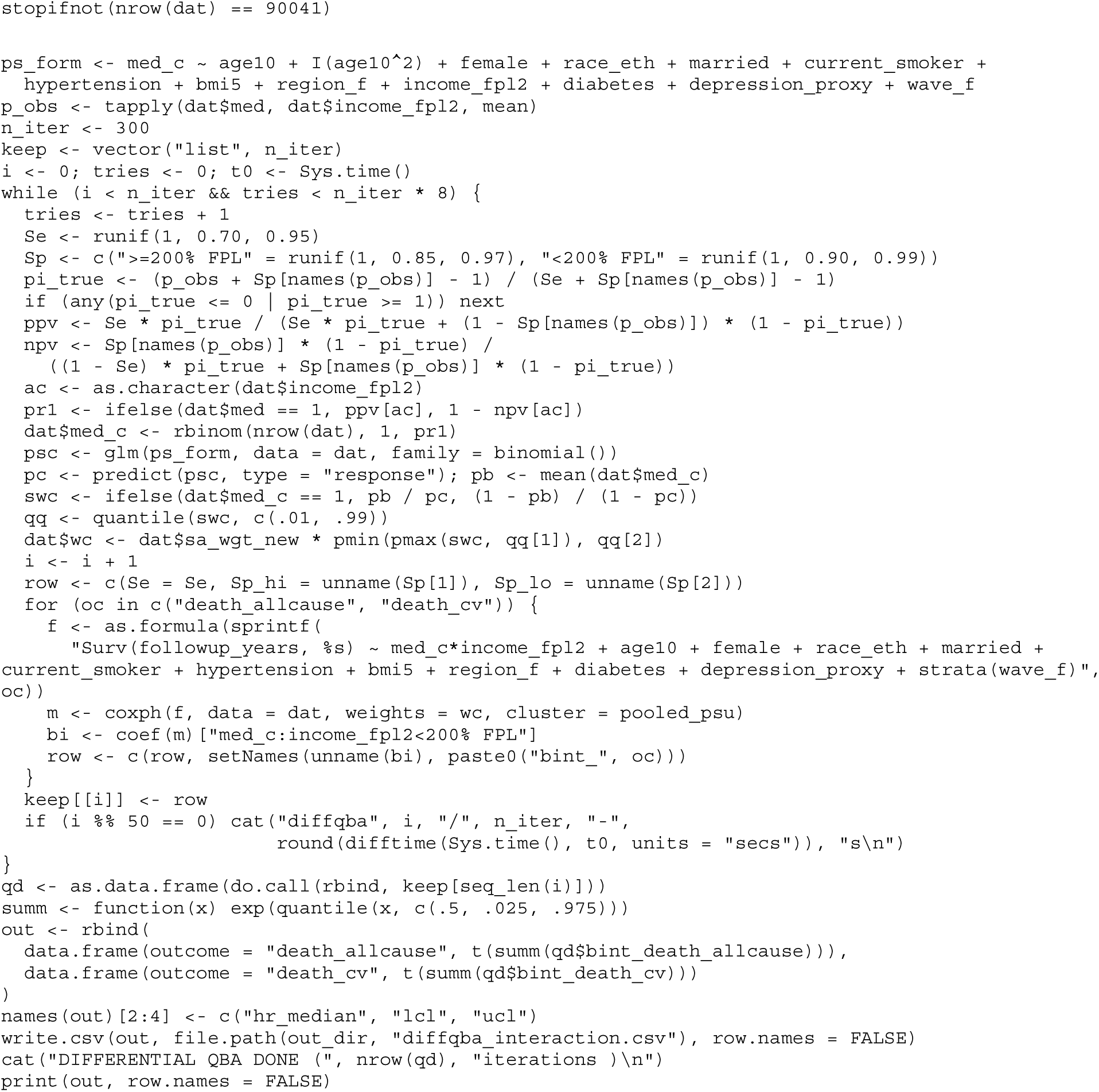

## Notes

### Competing Interest Statement

The authors have declared no competing interest.

### Author Declarations

The study used (or will use) ONLY openly available human data that were originally located at National Center of Health Statistics.

